# Exploring human mixing patterns based on time use and social contact data and their implications for infectious disease transmission models

**DOI:** 10.1101/2022.01.25.22269385

**Authors:** Thang Van Hoang, Lander Willem, Pietro Coletti, Kim Van Kerckhove, Joeri Minnen, Philippe Beutels, Niel Hens

## Abstract

**Background:** The increasing availability of data on social contact patterns and time use provides invaluable information for studying transmission dynamics of infectious diseases. Social contact data provide information on the interaction of people in a population whereas the value of time use data lies in the quantification of exposure patterns. Both have been used as proxies for transmission risks within in a population and the combination of both sources has led to investigate which kind of social encounters are most relevant to describe transmission risk.

**Methods:** We used social contact and time use data from 1707 participants from a survey conducted in Flanders, Belgium in 2010-2011. We calculated weighted exposure time and social contact matrices to analyze age- and gender-specific mixing patterns and to quantify behavioral changes by distance from home. We compared the value of both data sources, individually and combined, for explaining seroprevalence and incidence data on parvovirus-B19, Varicella-Zoster virus (VZV) and influenza-like illnesses (ILI), respectively.

**Results:** Assortative mixing and inter-generational interaction is more pronounced in the exposure matrix due to the high proportion of time spent at home. This pattern is less pronounced in the social contact matrix, which is more impacted by the reported contacts at school and work. The average number of contacts declined with distance, however on the individual-level, we observed an increase in the number of contacts and the transmission potential by distance when travelling.

We found that both social contact data and time use data provide a good match with the seroprevalence and incidence data at hand. When comparing the use of different combinations of both data sources, we found that the social contact matrix based on close contacts of at least 4 hours appeared to be the best proxy for parvovirus-B19 transmission. Social contacts and exposure time were both on their own able to explain VZV seroprevalence data though combining both scored best. Compared with the contact approach, the time use approach provided the better fit to the ILI incidence data.

**Conclusions:** Our work emphasises the common and complementary value of time use and social contact data for analysing mixing behavior and infectious disease transmission. We derived spatial, temporal, age-, gender- and distance-specific mixing patterns, which are informative for future modelling studies.

## Background

Infectious diseases have substantial impact on public health and economy and warrant constant monitoring and follow-up. Initially, disease transmission models relied on untested assumptions about “at risk events”. In recent years, the models have been informed by data from social contact surveys such as POLYMOD [1] in which participants had to report about their contact behavior by age, gender, frequency, etc. Social contact patterns have been successfully used as proxies for transmission of close-contact diseases, such as influenza and mumps, under the so called *social contact hypothesis* [2]. The use of social contact data helps estimating key epidemiological parameters [3, 4], behavioral changes [5, 6, 7] and demographic change [8] in the context of disease transmission. The number of social contact surveys to collect empirical data on human contact behavior has increased substantially over recent years [9].

Time use data have also proven their value for explaining infectious disease data using the *time use approach* in which mixing patterns can be estimated from the time spent at a given location [10, 11, 12]. The reported presence over time at different locations during a day enables the estimation of the exposure time among age groups [12]. The notion of co-presence is complementary to reported social contacts, hence time use data are useful to capture “at risk events” that fall outside the definition of a social contact.

The integration of both the “time use approach” and the “social contact approach” has lead to estimation of “suitable contacts” [11]. De Cao et al. [11] showed that the interplay between exposure time and social contacts appeared to be important to study the transmission of Varicella-Zoster virus (VZV) whereas the transmission dynamics of parvovirus-B19 was best captured using only exposure time. This study was based on two independent surveys.

Contact dispersal is of essence to capture disease transmission dynamics, though a systematic review of social contacts surveys [9] revealed that only a limited number of contact studies investigated the relationship between contacts and distance. Disease counts have been modeled using power law dispersal kernels [13, 14, 15], though questions remain whether this also holds for social mixing behavior. A study in Great Britain [16] collected information on the distance from home for each contact and observed a decrease in contact duration with increasing distance from home. In addition, age-specific contact patterns and temporal differences with respect to weekdays and weekends have been described [9], but gender-specific behavior in social mixing is less reported, though could contribute to the parameterisation of mathematical transmission models [17, 18, 19].

In this work, we use data from a comprehensive survey to compare the social contact, time use and suitable contact approach. An important element in this survey [19] is the recording of the distance from home for every reported location in the time use survey. In particular, we (1) compared mixing patterns based on time use and social contact data, and explored covariates such as age, gender, location, etc.; (2) analysed social mixing patterns and estimated basic reproduction numbers by distance from home; (3) evaluated the value of time use and social contact data to explain seroprevalence data for VZV and parvovirus-B19, and influenza-like illness (ILI) incidence data in Belgium.

## Methodology

This analysis is based on three types of data: social contacts, time use, and clinical information w.r.t. respiratory diseases. We first introduce each data set and continue with the methods to handle missing data and uncertainty. Next, we describe how we analysed exposure time and social contact data and calculated mixing matrices. We end with the integration and evaluation of different mixing patterns in epidemiological models for VZV, parvovirus-B19 and ILI incidence data from Belgium.

### Data

This study is based on a diary-based survey on social contacts and time use, which was conducted between September 2010 and February 2011 in Flanders, Belgium. The general description of this data can be found in [19] and the dataset is available online within the social contact data sharing initiative [20] and the SOCRATES platform [21]. The final sample size used for the analysis in this work is 1707 participants, as in [19].

To record time use, participants were asked to indicate for pre-defined time slots the location at which they spent most of their time. Location types were pre-defined (home, kindergarden, school, workplace, transport, family, leisure and other) and the options were adapted to the age of the participant. For example “kindergarden” was only used for surveying children and “workplace” only for adults. We also questioned the distance from home for each location in four categories (0-1 km, 2-9 km, 10-74 km or 75 km or more). Time slots were mostly of 1-hour length, except in the morning (2-5h, 5-8h) and in the evening (20-22h, 22-24h, 24h-2h). With respect to missing data, 1486 participants (87%) provided full information on their location and distance from home for each time-slot, and 49 participants provided no time use data at all.

Seroprevalence data was used from the Belgian sero-survey in 2001-2003 on parvovirus-B19 and VZV. In total, 3080 sera were tested for parvovirus-B19 and 3256 sera were tested for VZV, from which 2975 sera were tested for both VZV and parvovirus-B19. The sero-survey involved participants ranging from 0 to 71.5 years of age. More information is provided in Hens et al [22].

The ILI incidence data were collected from a network of general practitioners (GPs) in Belgium. The GPs in the network were asked to report the weekly number of ILI by 7 age groups: *<*1, 1-4, 5-14, 15-19, 20-64, 65-84, ≥ 85 years. The data were provided by the Scientific Institute of Public health. More details of the data are described in [23] and [24].

### Analysis

#### Missing data

Missing values in the time use data were treated as “Missing at Random” [25]. As such, we assumed a systematic relationship between the propensity of missing values and the observed data, but not with the missing data. Missing data were imputed by using Multivariate Imputation via Chained Equations (MICE) [26]. The list of variables included in the imputation model can be found in Additional file 1 Table S3. By applying MICE with different random number generator seeds, we obtained 10 imputed data sets. Please note that for the initial data exploration (Table 1 and Appendix), as described in the first part of the Results section, we used the original data with location type set to “Missing”.

**Table 1:** Reported daily time use by location with respect to age, gender, type of day, period, health state, household type.

| Covariate | N | Home (%) | School (%) | Work (%) | Transport (%) | Other (%) | Missing (%) |
| --- | --- | --- | --- | --- | --- | --- | --- |
| **Age** | **1707** |  |  |  |  |  |  |
| [0-3] | 87 (5.10%) | 77.82 | 8.53 | 0 | 2.06 | 9.44 | 2.15 |
| [3-6] | 84 (4.92%) | 74.51 | 11.96 | 0 | 2.88 | 9.33 | 1.32 |
| [6-12] | 125 (7.32%) | 64.50 | 15.30 | 0 | 2.60 | 13.24 | 4.36 |
| [12-18] | 79 (4.63%) | 64.72 | 11.66 | 1.11 | 4.43 | 15.88 | 2.20 |
| [18-25] | 99 (5.80%) | 58.67 | 7.45 | 9.47 | 4.71 | 16.46 | 3.24 |
| [25-45] | 524 (30.70%) | 61.86 | 0.29 | 16.9 | 4.18 | 13.19 | 3.58 |
| [45-65] | 468 (27.42%) | 62.36 | 0.20 | 12.77 | 4.18 | 13.79 | 6.70 |
| 65+ | 241 (14.11%) | 76.31 | 0 | 0.48 | 4.26 | 11.50 | 7.45 |
| **Gender** | **1707** |  |  |  |  |  |  |
| Female | 917 (53.72%) | 66.34 | 2.82 | 8.33 | 3.72 | 13.72 | 5.07 |
| Male | 790 (46.28%) | 64.78 | 3.77 | 10.54 | 4.21 | 12.28 | 4.42 |
| **Day type** | **1705** |  |  |  |  |  |  |
| Weekdays | 1297 (76.07%) | 64.09 | 4.24 | 11.71 | 3.98 | 11.52 | 4.46 |
| Weekends | 408 (23.93%) | 70.77 | 0.15 | 1.91 | 3.86 | 17.99 | 5.32 |
| **Periods** | **1705** |  |  |  |  |  |  |
| Regular | 1268 (74.37%) | 64.91 | 4.24 | 10.39 | 3.97 | 11.97 | 4.52 |
| Holiday | 437 (25.63%) | 67.97 | 0.43 | 6.39 | 3.89 | 16.25 | 5.07 |
| **Feeling ill** | **1700** |  |  |  |  |  |  |
| Yes | 45 (2.65%) | 86.39 | 0.83 | 0.28 | 1.85 | 8.42 | 2.23 |
| No | 1655 (97.35%) | 65.21 | 3.34 | 9.64 | 4.02 | 13.18 | 4.61 |
| **Family status** | **1008** |  |  |  |  |  |  |
| Living with children | 238 (23.61%) | 64.63 | 0.49 | 15.30 | 3.66 | 11.15 | 4.77 |
| Living without children | 770 (76.39%) | 61.44 | 0.17 | 14.53 | 4.35 | 14.21 | 5.30 |

#### Uncertainty

We applied a stratified bootstrap of the participant data (including social contact and time use data) by 5-year age categories to construct confidence intervals for the outcome. We created 1000 bootstrap replicates for each imputed data set (ℳ=10), resulting in 10000 data sets [27] (See Additional file 1 for more details).

#### Participant weights

Participant contributions to mixing patterns were weighted to account for sampling probabilities based on the joint distribution of age, household size and day type (holiday/regular periods and weekday/weekend). We used census data for Belgium from 2011 (http://epp.eurostat.ec.europa.eu) as reference population and weights were constrained to a maximum of 3 to reduce the impact of the corresponding observations.

#### Time use patterns

We explored time use patterns by calculating the proportion of time at each location as reported by participants. To limit data sparseness, we combined “kindergarten” with “school”, and also “family” and “leisure” with “other”. Reported proportions were analysed in relationship with age, gender, weekday/weekend, period (regular/holiday), health state (ill or not ill) and family status. The latter was only relevant for adults between 25 and 65 years of age and defined as living with or without children less than 13 years old. We used 7 age categories in line with the Belgian education and employment system; 0-2 years, 3-5 years, 6-11 years, 12-17 years, 18-44 years, 45-64 years and 65-90 years of age. We also performed an additional analysis of the reported locations using 5-year age groups to study gender-specific differences in time use (refer to Additional file 2 Figure 2).

The proportion of time spent at different locations during one day can be analysed as compositional data. This is a special type of multivariate data, with each proportion being non-negative and the total sum adding up to one. A divergence based regression modelling technique is used to explore time use at each location [28]. More specifically, we used Kullback-Leibler divergence to minimize the distance between the observed and the fitted compositions with respect to the coefficients of the regression (Additional file 3). We used the 10000 datasets generated through MICE and bootstrapping to construct weighted confidence intervals for all parameter estimates [29].

#### Contact patterns taking into account time use and distance

We used a GAMLSS model [30] for analysing the determinants of social contact patterns, while taking into account the time spent at each location as explanatory variables. To handle classification issues in this sub-analysis, we aggregated all location types into “home”, “work”, “school” and “other”.

We linked social contact and time use data to obtain for each social contact the time spent at the reported location and the distance from home. Participants were able to report multiple distances for one location type over different time-slots in the time use part. For example, one could report “other” at 2 km and 10 km from home, for shopping in the morning and sport activities in the evening. As such, we applied a probabilistic link procedure between a social contact and the distance from home, based on the relative time spent at each distance per location type. The interaction between travel and social contact patterns has been studied unconditionally and conditionally upon presence at each distance, i.e., the former presents the population average and the latter is in line with the individual-level perspective. To elaborate on social contact dispersal, we classified participants by age using a cutoff of 18 years (child and adult), gender and type of day (regular weekday, weekend, holiday). We excluded the contacts with missing distance (±7%) to calculate the weighted average number of contacts by distance and by age, gender and day type.

#### Social contact matrices and R_0_

The matrix **M**^*dt*^, representing the mean number of contacts at distance *d* during one day of type *t*, can be estimated by the following expression:

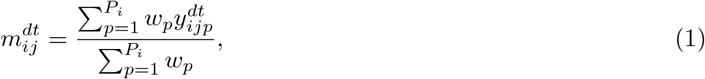

where *P*_*i*_ is the number of participants in age class *i, w*_*p*_ the weight for participant *p* and 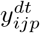 the reported number of contacts made by participant *p* of age class *i* with someone of age class *j* at distance *d* during one day of type *t*. The social contact matrix *c*_*ij*_, representing the per capita daily contact rate between age classes, was calculated as

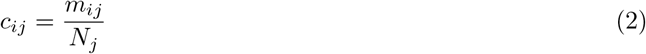

with *N*_*j*_ the population size in age class *j*, obtained from census data.

The next generation matrix *G* with elements *g*_*ij*_ indicates the average number of secondary infections in age class *i* through the introduction of a single infectious individual of age class *j* into a fully susceptible population [31]. Assuming a rectangular population age-distribution, the next generation matrix at distance *d* during one day of type *t* is defined by:

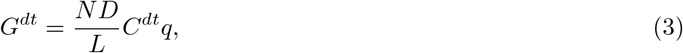

with *N* the population size, *D* the mean duration of infectiousness, *L* the life expectancy, *C*^*dt*^ the contact matrix at distance *d* during one day of type *t* and *q* the proportionality factor. The basic reproduction number R_0_ can be calculated as the dominant eigenvalue of the next generation matrix. To enable the comparison of transmission dynamics by distance, we needed to specify the disease-specific q parameter of equation (3). As such, we constrained the average *R*_0_ for regular weekdays using all reported contacts from the bootstrapped and imputed data sets to 2.

#### Exposure time matrices

We calculated the age-specific exposure times based on time use data following the Proportionate Mixing Assumption (PTM) as previously used by [10]. As such, for one single location and time slot of the survey day, we calculated the exposure time of a participant to the other participants proportionally to their relative participation in that location. We used 17 categories; 0-2 years, 3-5 years, 6-11 years, 12-17 years, 18-25 years, 5-year age categories between 25 and 80 years of age, and a closing category of 80-90 years of age. Participants had to report their household members in terms of age, gender and whether they were present at home during the survey day. This allowed us to compute the time of exposure between members of the same household, which is formalized in the matrix **H**.

For locations other than home, the exposure time between people in age group *i* and *j* at specific location *l* and time slot 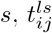, can be computed under the PTM assumption as follows:

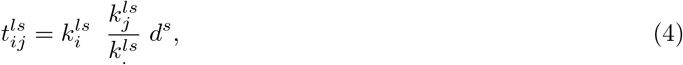

where 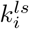 and 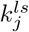 are the number of people present at location *l* during time slot *s* in age group *i* and *j*, respectively, 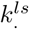 is the sum over all age classes at location *l* during time slot *s* and *d*^*s*^ is the duration of each time slot *s* in hours. From (4), we can compute the time of exposure between people in age group *i* and in age group *j*, referred to as matrix **T**, as follows:

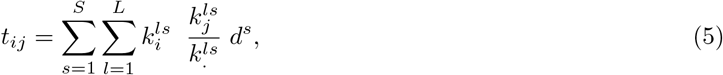

The sum of **T** and **H** determines the overall exposure time matrix **E**, with elements *e*_*ij*_:

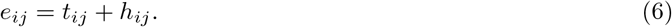

The response matrix **E** contains non-negative quantities that are considered to follow a mixed discrete-continuous distribution, comprising value *Y* = 0 with probability *p*_0_ and value *Y* = *Y*_1_ ∈ (0, ∞) with probability (1-*p*_0_). The R package gamlss.inf [32] was used to model **E**. For the discrete part (zero or not), we created a binary response variable to fit a binary logistic model. We explored different distributions for the continuous part (*Y >* 0) such as Gamma, Inverse Gaussian, Inverse Gamma, Log-normal, Weibull and Pareto. Participants weights were taken into account and model selection was based on the Akaike information criterion (AIC).

#### Suitable contact matrices

The “suitable contact” approach assumes that not all social contacts are informative for disease transmission and that long duration and more intimate contacts are more likely to be relevant. To construct suitable contact matrices, we followed the procedure of De Cao et al. [11], which considered contacts and exposure between age classes *i* and *j* (*c*_*ij*_ and *e*_*ij*_, respectively). Let *u*_*ij*_ be a random variable representing the number of suitable contacts between age group *i* and *j*. Then the expected number of suitable contacts is given by the product of the average number of contacts *c*_*ij*_ and the proportion of these contacts that are relevant for transmission 1 − exp(−*e*_*ij*_*/c*_*ij*_) where exp(−*e*_*ij*_*/c*_*ij*_) is the Poisson probability that a contact is not suitable. Therefore the probability of infection *β*_*i,j*_ is given by

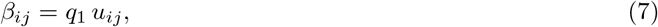

where *q*_1_ is a constant disease-specific transmission coefficient and

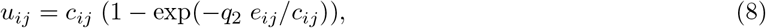

with *q*_2_ the fraction of total exposure time between age groups that is relevant for transmission.

#### Fitting mixing matrices to serological data for parvovirus-B19 and VZV

The social contact and the time use approach rely on the assumption that infectious disease transmission between people in different age categories is proportional to their number and duration of their physical encounters, respectively. To complement the age-specific social contact and exposure time matrices within a transmission model framework, we estimated *q*_1_ and *q*_2_ from Equation 7 and 8 to capture the reported disease prevalence under endemic equilibrium. We used a transmission model for VZV based on Maternally-derived immunity, Susceptible, Infectious and Recovered infection states (MSIR) [3, 11, 33]. For parvovirus-B19, we used an MSIRWb model [33], which is an MSIR model augmented with waning immunity and boosting in the Recovered compartment. In both models, and without loss of generality, we assumed newborns to be fully protected by maternal antibodies until 6 months of age [33], after which they become susceptible.

#### Fitting mixing matrices to influenza-like illnesses (ILI) incidence data

We used an age-structured Susceptible-Exposed-Infectious-Recovered (SEIR) model with different mixing matrices to capture the Belgian ILI incidence in 2010-2011. The weekly ILI incidence was recorded by the general practitioners (GPs) network, which covered 1.75% of the population in Belgium [23, 24]. Because we only considered one season (52 weeks from week 40 in 2010 to week 39 in 2011), newborns and mortality are not included in the model (see Additional file 5 for more details of the model description and parameters). We used an average latent period of 1 day and an average infectious period of 3.8 days based on the literature [34, 35]. We estimated the model parameters by minimizing the sum of squared differences between the observed ILI incidence rate and the predicted incidence rates. The following parameters were estimated: age-specific and proportionality factors *q*_*i*_ based on the social contact hypothesis [2] and a scaling factor. The scaling factor is used to align the predicted incidence rates to the observed incidence rates, which accounts for those individuals with influenza who do not seek medical attention (the consultation rate) [34, 35, 36].

## Results

### Time use patterns

The final sample size for the analysis is 1707 participants, as reported in [19], though 49 participants left the time use part entirely blank. People older than 45 years of age accounted for the highest proportion of missing data for at least one time slot (Table 1). People spent on average about two thirds of their day at home.

The elderly over 65 years of age and children less than 6 years old reported the highest proportion of time spent at home. For the active population, we observed that time at work is higher for males than for females, hence the opposite is observed for time spent at home. Temporal factors such as day of the week and holiday periods had a large impact on time use.

Health status was also linked to substantial changes in participants’ time use. Participants did report an increase of 5 hours at home when feeling ill and almost no time at work. However, the time spent at other locations is still substantial when people reported feeling ill. Adults between 25 to 65 years of age living with children under 13 years of age reported more time spent at home compared to those without children.

We analysed the reported time at work more in detail and observed that the average time is similar for males and females until the age of 30 years (Additional file 2 Figure S1). Differences emerge from the age of 30 years, in which females reported on average less time at work compared to males in the same age group. We observed the highest differences in age groups [40,45), [50,55) and [55,60) in which males reported 10% more time at work compared to females. The reported time at work declined after 65 years of age for both genders. The time spent at “other” places was similar between males and females in general, except for the age categories [6,12) and [18,25) in which males reported 6% more compared to females. The reported time for transport was similar for males and females.

We analysed the time use patterns with a divergence-based regression model with multinomial logit link function including gender, age, day type and period. Table 2 shows all parameter estimates and 95% confidence intervals. After controlling for age and temporal factors, time at work reported by males is significantly higher compared to females. In reverse, females reported more time at home than males. The gender-specific difference in time spent at school was not statistically significant. Age had a significant effect on presence at home, school and work but not on the reported time at location category “other”. As expected, temporal factors played a crucial role in the time use patterns, particularly on the presence at work and school. We also observed a clear increase in the reported time in the “other” category during weekends vs. regular weekdays. This denotes compensation behavior of people not at work or at school, which needs to be considered when modeling weekend days.

**Table 2:** Parameter estimates and 95% confidence intervals of the divergence-based regression analyses for location-specific time use patterns. The asterisks (*) denote the confidence intervals which do not include zero.

| Variable | Category | Home | School | Work | Other places |
| --- | --- | --- | --- | --- | --- |
| **Intercept** |  | 2.72 [2.40;3.10]* | 1.62 [1.22;2.03]* | -1.40 [-9.16;-0.37]* | 1.18 [0.80;1.55]* |
| **Gender** | Male | -0.17 [-0.35;-0.03]* | -0.14 [-0.41;0.10] | 0.37 [0.13;0.57]* | -0.19 [-0.4;-0.01]* |
| *Ref.: Female* |  |  |  |  |  |
| **Age** | [0-3] years | 1.05 [0.55;1.64]* | 0.44 [-0.23;1.14] | -7.35 [-51.28;-2.61]* | 0.30 [-0.29;0.97] |
| *Ref.: [12-18] years* | [3-6] years | 0.55 [0.08;1.13]* | 0.44 [-0.08;1.09] | -7.38 [-40.00;-0.77]* | -0.17 [-0.70;0.46] |
|  | [6-12] years | 0.52 [0.05;1.03]* | 0.74 [0.24;1.29]* | -7.5 6 [-44.18;-1.99]* | 0.34 [-0.12;0.89] |
|  | [18-25] years | -0.04 [-0.5;0.42] | -0.53 [-1.10;-0.01]* | 2.20 [1.09;9.82]* | 0.06 [-0.43;0.56] |
|  | [25-45] years | 0.01 [-0.36;0.37] | -3.74 [-4.53;-3.09]* | 2.94 [1.93;10.71]* | -0.13 [-0.51;0.27] |
|  | [45-65] years | 0.09 [-0.31;0.44] | -4.17 [-5.14;-3.48]* | 2.55 [1.52;10.24]* | -0.04 [-0.42;0.35] |
|  | 65+ years | 0.17 [-0.22;0.58] | -7.81 [-22.25;-4.49]* | -0.93 [-2.57;6.46]* | -0.35 [-0.74;0.09] |
| **Temporal factor** | Holiday weekday | 0.4E-1 [-0.23;0.23] | -2.48 [-3.52;-1.84]* | -0.24 [-0.56;0.07] | 0.26 [0.02;0.53]* |
| *Ref.: Regular weekday* | Regular weekend | 0.11 [-0.09;0.35] | -3.36 [-5.15;-2.55]* | -1.92 [-2.56;-1.42]* | 0.54 [0.29;0.81]* |
|  | Holiday weekend | 0.23 [-0.09;0.55] | -5.34 [-19.6;-4.09]* | -1.90 [-2.69;-1.32]* | 0.60 [0.27;0.94]* |

### Social contacts patterns taking into account time use

We analysed the number of reported social contacts with a zero-inflated negative binomial model and observed that the overall number of contacts was inversely associated with the time spent at home, and positively associated with the time spent at school or work (Table 3). The number of contacts at school or work tends to increase with the time spent in these settings. We observed a gender effect, implying that males tend to have on average fewer contacts compared to females per time-unit. However, there was no statistical difference in the number of contacts at different locations between males and females. With respect to age, the highest overall number of contacts were observed among children up to 18 years of age and among people in the working age (25-65 years of age). The age effect was not observed for the social contacts at home and school, but observed for the social contacts at other places.

**Table 3:**
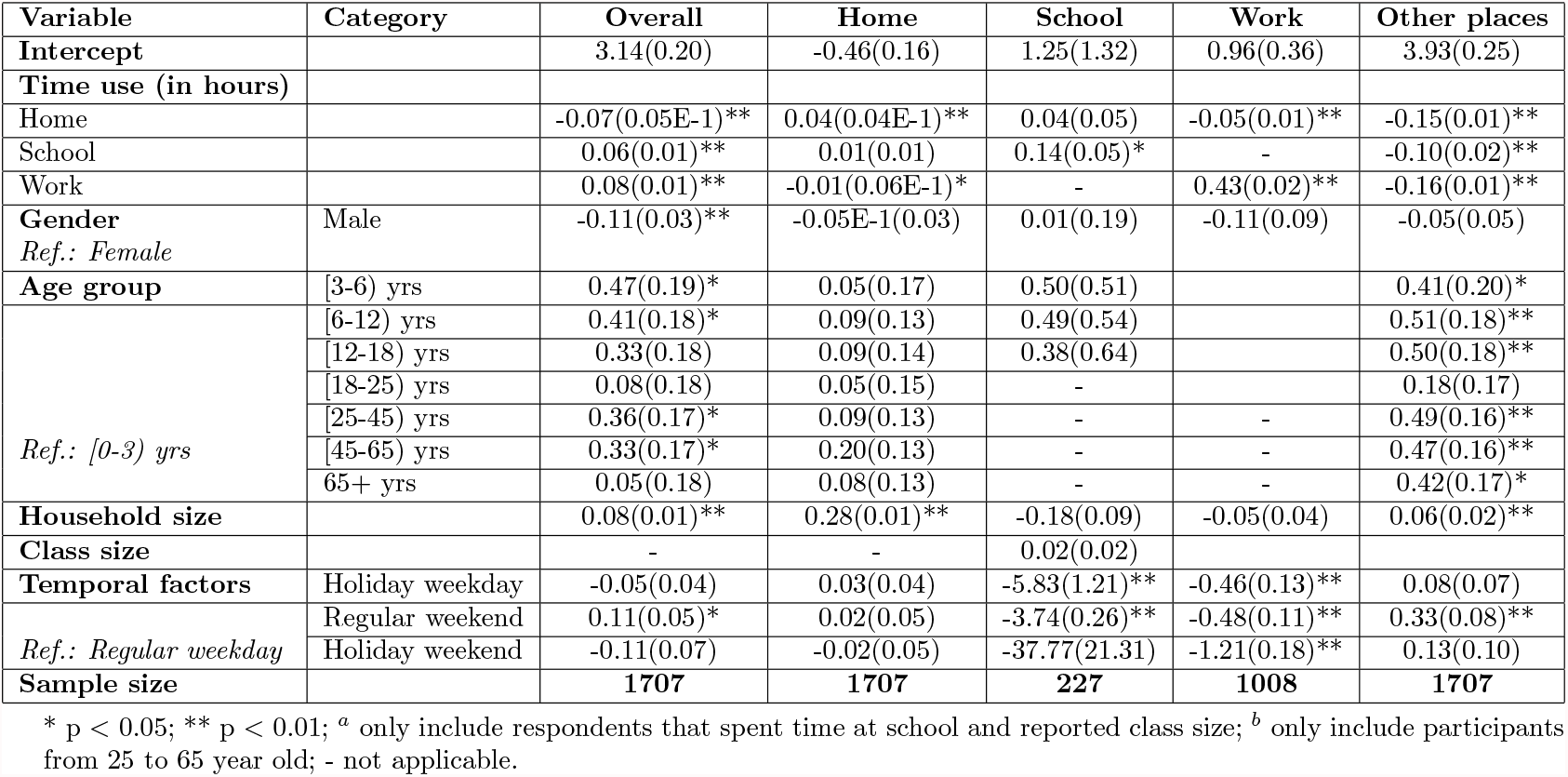
Parameter estimates and p-values of zero-inflated negative binomial models for the total number and location-specific social contacts using age, gender, temporal and time use covariates.

Household size was positively associated with the total number of contacts, contacts at home and other places, however household size showed no significant effect on the social contacts at school and at work. The reported class size did not have a clear effect on the number of school contacts. As expected, we observed an explicit link between temporal factors and the number of school and work contacts.

### Social contact dispersal

To explore social contact dispersal, we analyzed the social contact data by distance in combination with age, gender and type of day (Figure 1). At population level, i.e. unconditional upon whether or not people travel, we observed most contacts for children (0-18 years of age) in the category 0-9 km from home during weekdays and very few contacts beyond 10 km from home. This pattern was the same for boys or girls. During weekends, children reported a decline in the number of contacts by distance, but less so for girls. For adults (18+ years of age), we observed an increase by distance on weekdays up to 10-75 km from home. On average, people reported almost no contacts 75+ km from home. Females reported more contacts up to 9 km from home compared to males, though the opposite holds for the category 10-74 km. For weekends, we observed a decline in the number of contacts beyond 10 km from home, both for males and females.

**Figure 1:**
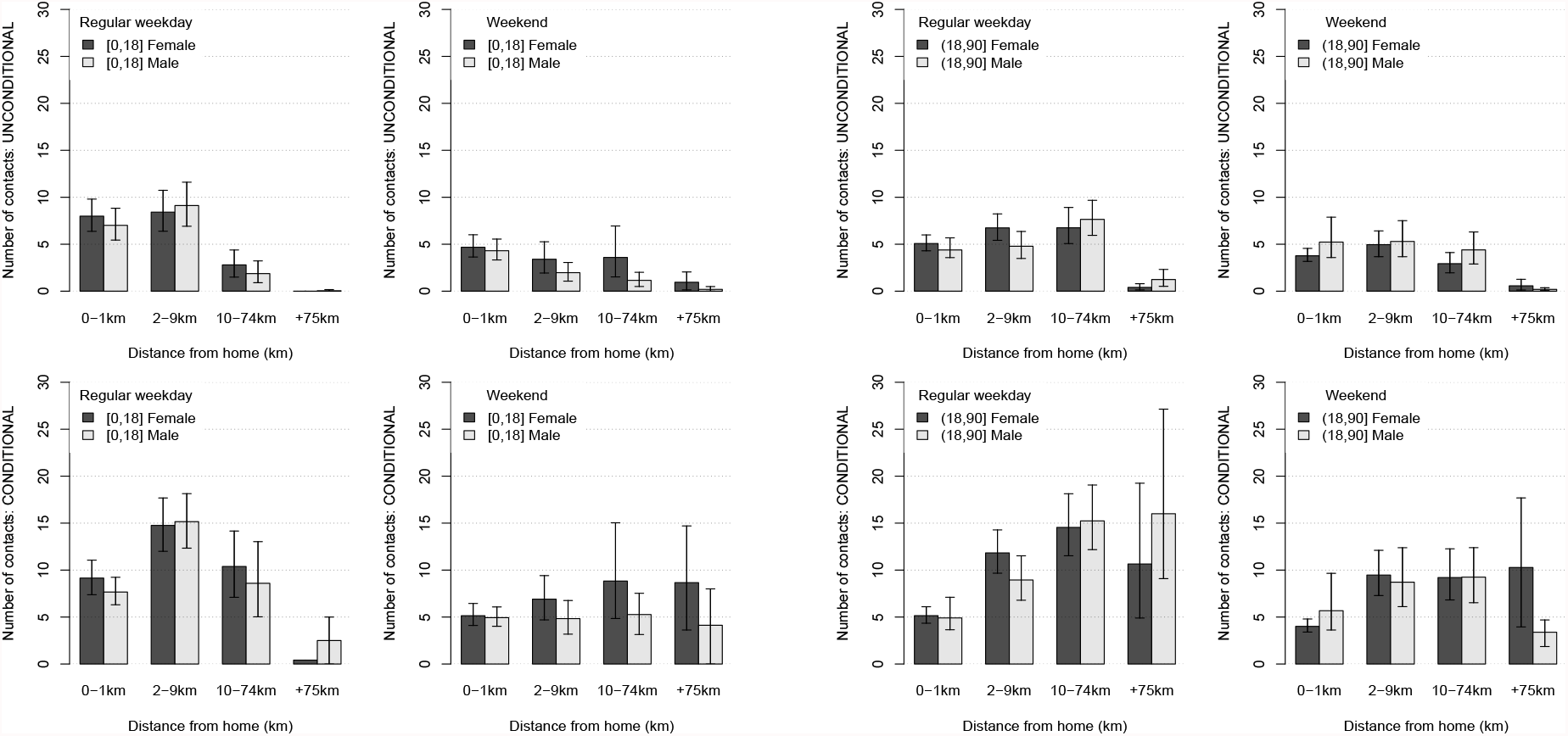
Number of contacts by distance unconditional (top) and conditional (bottom) upon presence by age (left/right) and gender (color) during regular weekdays and weekends. The median is represented by the bars, 95% CI by the whiskers.

The conditional results, i.e. looking at the number of contacts upon having traveled a certain distance, showed a different result compared to the unconditional (or population based) results. For children, the conditional results for weekdays show a clear maximum in the category 2-9 km from home. The number of contacts at 10-74 km from home equals the number of contacts close to home. During weekends, the reported number of contacts for girls increased with distance though the social contact behavior for boys seemed indifferent with distance up to 74 km. For adults, the number of social contacts during weekdays at 2-9 km and 10-74 km from home was two and three times the number of reported contacts at home, respectively. Also for the last distance category (75+ km from home), we observed an increase compared to the contacts at home but with large uncertainty. If adults leave home during weekends, the reported number of contacts by adults seems indifferent to the distance up to 74 km. Only at 75+ km from home, females reported fewer social contacts, compared to males. In conclusion, the number of contacts decreased by distance at population level, though if people made the effort to go somewhere, they made it count in terms of social encounters.

### Transmission dispersal and R_0_

The social contact dispersal as illustrated in Figure 1 induces distinct transmission potential by distance. We calculated social contact matrices by distance for child/adult interactions for each bootstrapped data set (e.g., Additional file 4 Figure S4 and S5) and corresponding R_0_ values. We calibrated the disease-specific proportionally factor *q* for a flu-like disease with median R_0_ = 2 with the full set of social contact data from regular weekdays. The successive R_0_ values using distance-specific contact data should be interpreted as the relative transmission potential and are presented in Figure 2. On the population level, the transmission potential slightly increased until 74 km from home during regular weekdays. This can be explained by the strong assortative mixing 2-9 km from home and the work-related mixing between adults 10-74 km from home (Additional file 4 Figure S4). The overall reduction in the transmission potential during weekends, can be explained by the fewer number of contacts for the distance category 10-74 km from home. Social contacts at 75+ km from home had almost no impact on the unconditional transmission potential.

**Figure 2:**
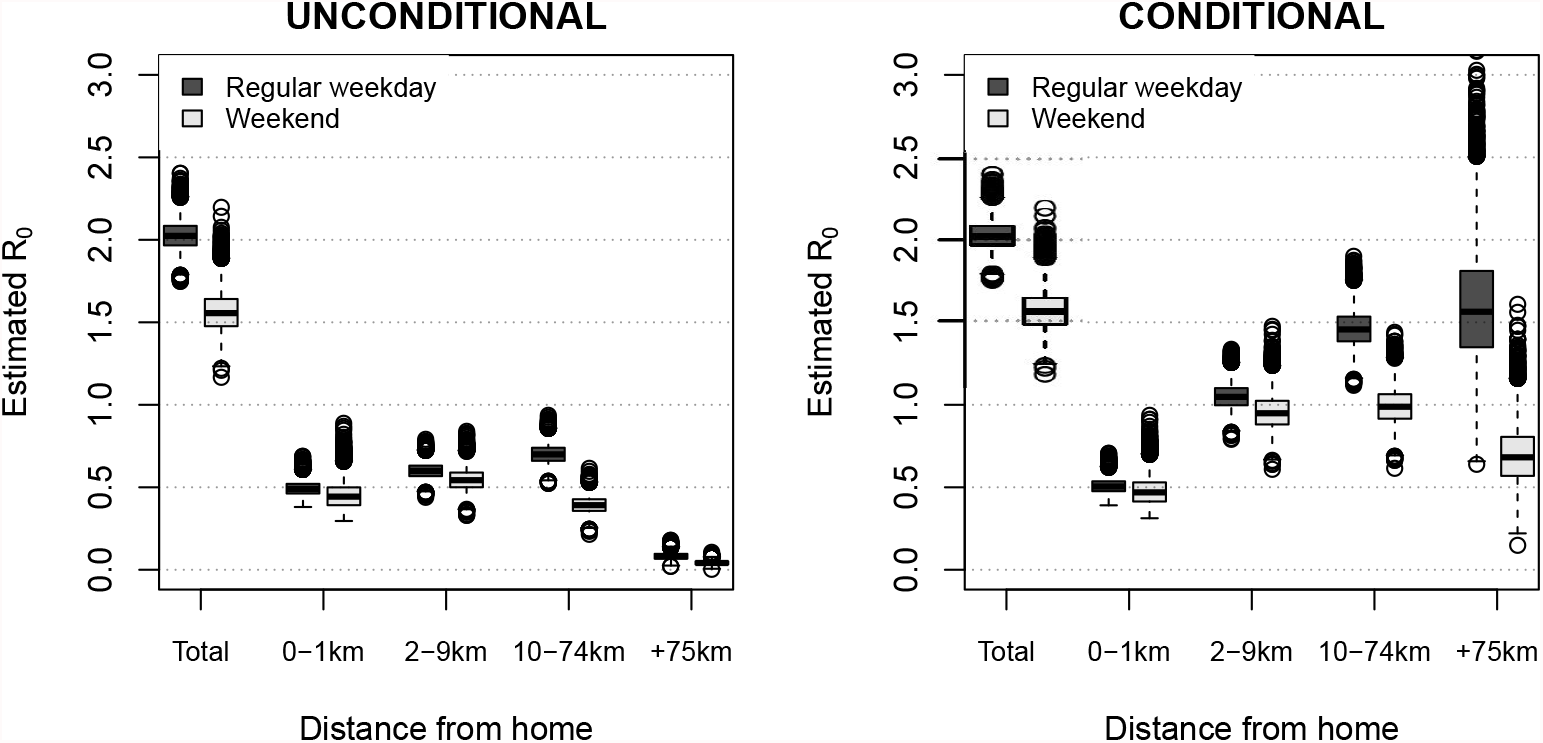
Estimated basic reproduction number R_0_ by distance, calculated as the leading eigenvector of the distance-specific social contact matrix and calibrated so the median R_0_ for regular weekdays equals 2 based on population-based (unconditional) contact matrices. For each distance category, we used the unconditional and conditional social contact patterns. The median is represented by the horizontal line in the box (75% interval), the whiskers denote the 95% confidence intervals and the dots are outliers.

The transmission potential conditional upon presence showed a clear increase with distance from home during regular weekdays (Figure 2). This effect was similar for weekends, however with a decrease of the estimated R_0_ for the last distance category. In general, the estimated transmission potential increased by distance from an individual-based perspective.

### Exposure time and social contact matrices

We used a zero-adjusted log-normal model on the time use data to estimate the age-specific exposure matrices. The resulting exposure matrix reflected contributions from exposure at home, school, work, and other locations (Figure 3). The main diagonal indicates that people tend to spend time with people of similar age. The two sub-diagonals represent the mixing pattern between generations. We also calculated the corresponding social contact matrix representing the weighted average number of contacts by age. Both matrices showed strong assortative mixing by age, especially for young children and teenagers. Especially the exposure matrix showed strong interaction among family members, as a result of the reported time at home. This pattern is very clear in the location-specific exposure matrix at home (Additional file 3 Figure S1).

**Figure 3:**
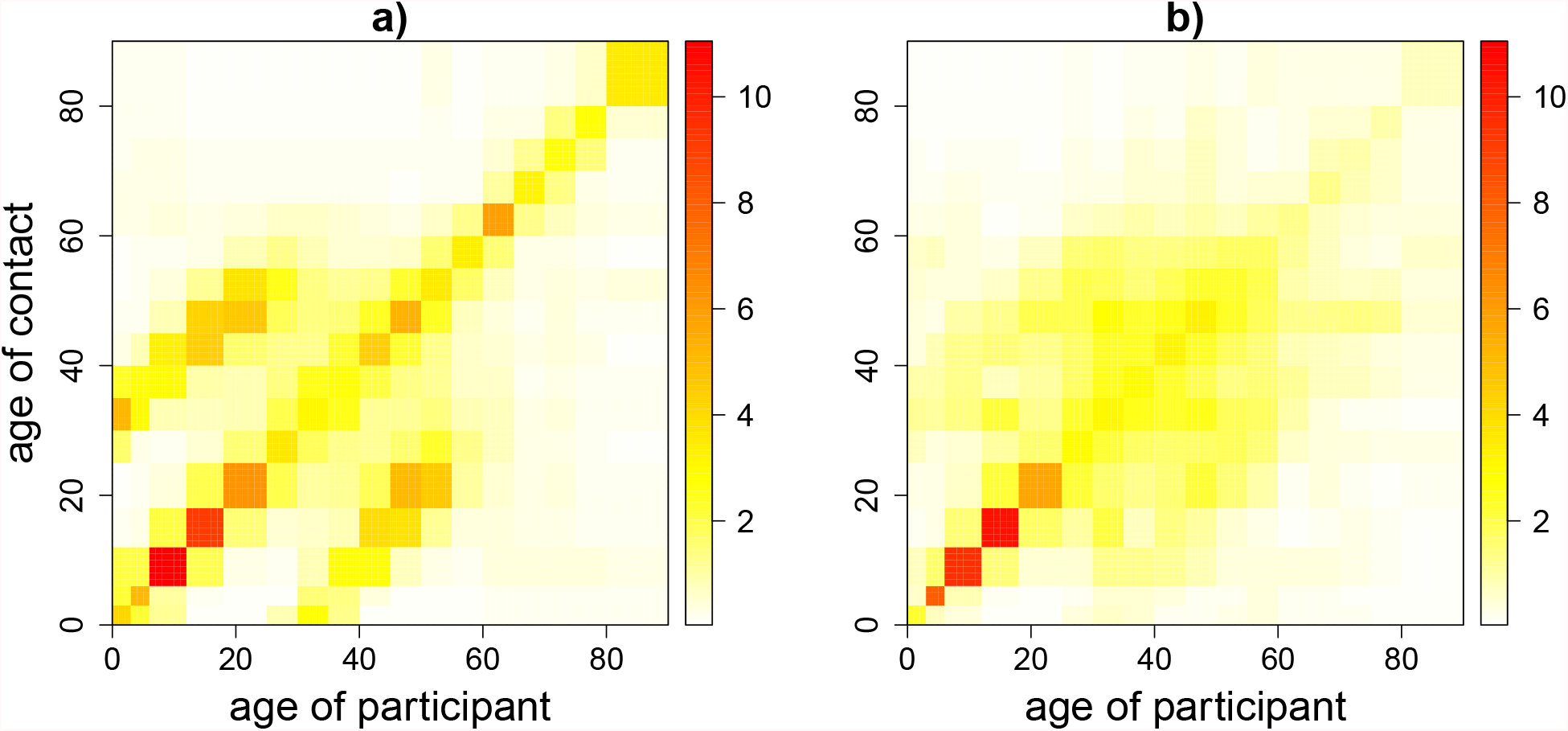
Mixing patterns by age represented by (a) weighted exposure time in hours and (b) weighted average number of contacts.

Time reported at home constituted on average up to ±66% of the total time per day, while contacts at home only accounted for ±18% of the total number of contacts. In contrast, the impact of employment is higher for the contact matrix compared to the exposure matrix (±40% of the total number of contacts though only ±9% of total time per day). We observed more pronounced assortative mixing pattern in the same-gender matrices for children and teenagers (Figure 4), though more different-gender exposure time for adults older than 50 years of age. In addition, the interaction between mothers and daughters seemed more pronounced compared to other child-adult interactions.

**Figure 4:**
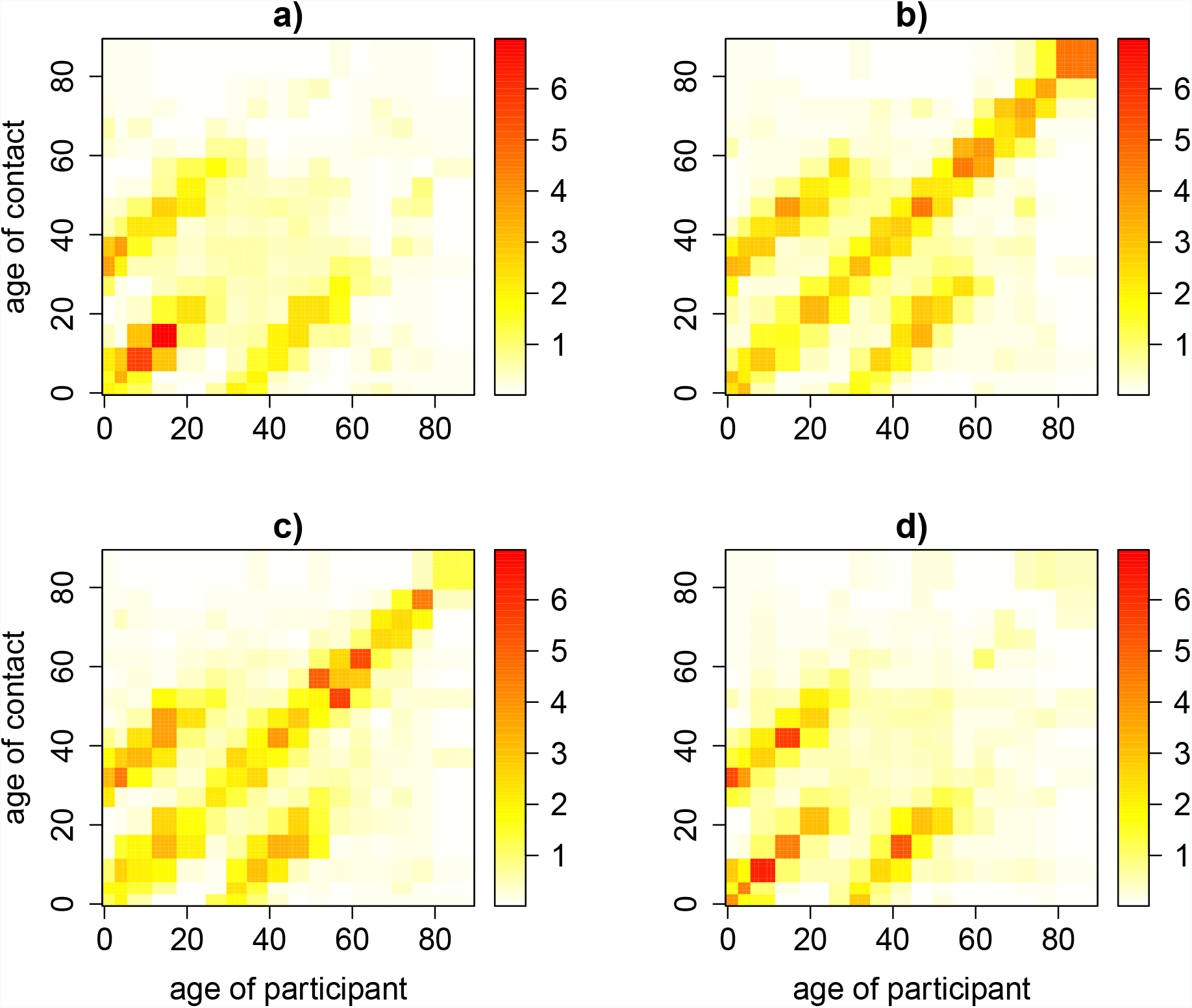
Estimated age- and gender-specific exposure time: (a)male-male, (b) male-female, (c) female-male and (d) female-female. The color scale indicates the exposure time in hours from low (white) to high (red).

### Fitting social contact and exposure matrices to parvovirus-B19 and VZV serological data

We tested the value of exposure and social contact matrices as a proxy for effective contacts governing transmission by their value in MSIRWb and MSIR models for parvovirus-B19 and VZV, respectively. Model estimations were compared to serological data and model selection was based on the AIC criterion. The overall time use matrix implied a better fit for both parvovirus-B19 and VZV compared to total social contact matrix. When exploring different levels of intimacy, location and duration, the social contact matrix based on close contacts lasting more than 4 hours, provided the best proxy for the transmission dynamics of parvovirus-B19 (Additional file 5 Table S2 and S3). For VZV, the social contact matrix based on close contacts of at least 1 hour did improve the contact approach, though the AIC was still higher compared to the time use approach.

The suitable contact approach, which accounts for both the number of social contacts and exposure time, scored slightly better for parvovirus-B19 compared to the contact approach, but not better in comparison to the time use approach. For VZV, the application of suitable contact estimates provided the overall best fit with the lowest AIC. The estimated proportionality factor *q*_2_ of exposure data with respect to the overall model prediction is much higher for VZV (0.94) than for parvovirus-B19 (0.13), which implied a higher number of suitable contacts for transmission of VZV. Our best-fitting models estimated a reproduction number of 1.9 [1.7; 2.1] and 7.8 [6.8; 8.5] for parvovirus-B19 and VZV, respectively. The estimated prevalence and force of infection by the 3 approaches were quite similar (Figure 5). We did observe differences in the predicted age-specific relative incidence by the time use and social contact approaches (Figure 6). The highest relative incidence obtained by the social contact approach was among children in the age group [12,18), while the highest relative incidence obtained by the time use approach is among children of [6-12) years. The latter also predicts a relatively higher incidence in adults between 35 and 50 years of age.

**Figure 5:**
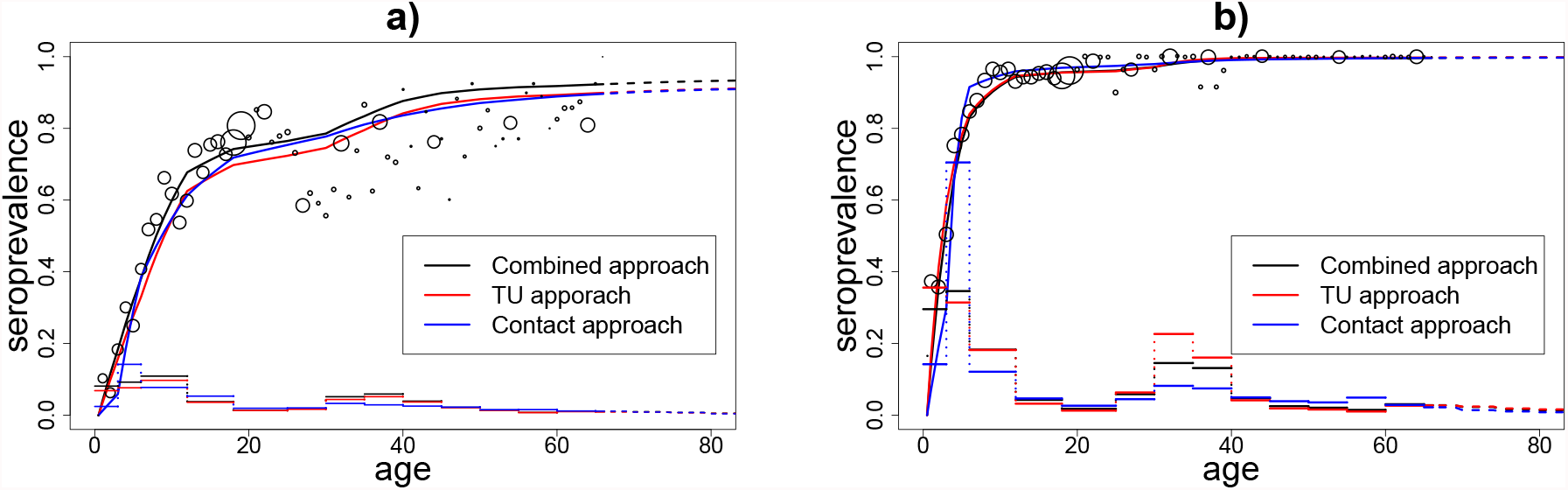
The fit of 3 different matrices to Belgian (a) serological parvovirus-B19 and (b) VZV data.

**Figure 6:**
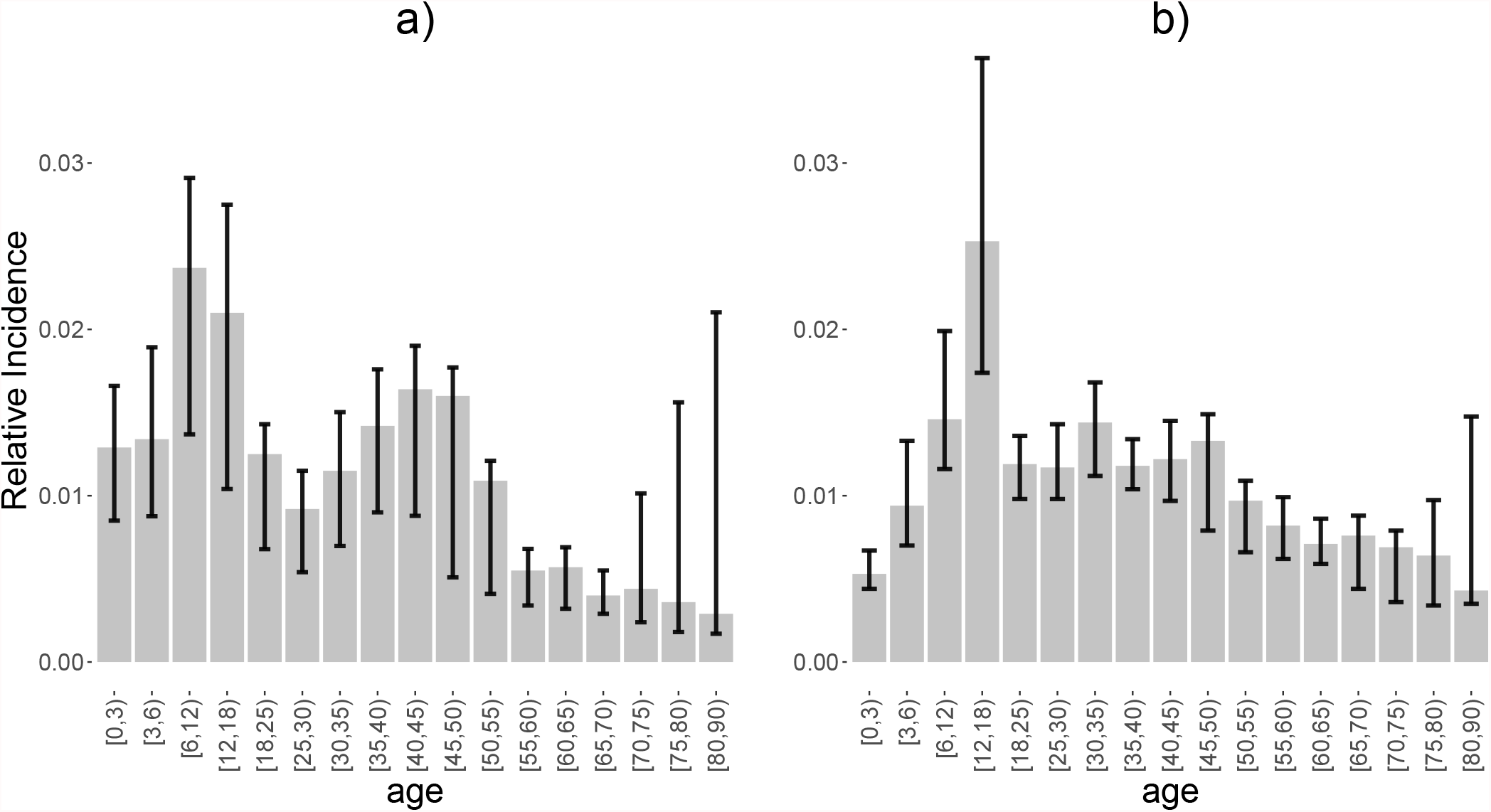
Estimated relative incidence by age based on (a) time use data and (b) social contact data.

### Fitting social contact and exposure time matrices to ILI incidence data

Social contact and exposure time matrices were used to compute transmission rates in the dynamic, differential-equation SEIR model for ILI incidence data. The model comparison was based on the least square score, a direct measure of goodness of fit with smaller values indicating a better fit to the ILI incidence data. The intimacy and duration of contacts seemed to matter for the ILI transmission modelling, such that physical contacts and contacts lasting at least 1 hour provided most information regarding transmission dynamics. The exposure time matrices provided a better fit for ILI compared to the total contact matrix (Additional file 5 Table S4). *R*_0_ estimated from the Based on the best scoring model, we estimated the *R*_0_ to be 1.43 with scaling factor of 0.266. The combination of contact and exposure time matrices did not improve the fit. Figure S1 (Additional file 5) shows the fit of different matrices to ILI incidence rate. The estimated incidence rates from models using suitable contact matrices and exposure time matrices are almost overlapping, with only a slight difference at the seasonal peaks of transmission.

Figure 7 shows the fit of the models based on the contact and exposure time matrices for the total population and different age groups. For each (sub)model, we present the least square value, the observed number and modeled based estimates of ILI cases, their ratio, and the mean absolute error (MAE). For the total population, the estimated ILI cases captures quite well the weekly observed number of ILI cases. The contact matrices and exposure time matrices provided quite similar results for the total population and most of age groups, except for the age group of 65 years and older. More precisely, the use of exposure time captured the observed ILI curve for this age group better compared to models informed by all social contact data. In general, the quality of the fit does not differ substantially between age groups. The models tend to overestimate the number of ILI cases for children and teenagers aged 0–19 years and adults aged 20–64.

**Figure 7:**
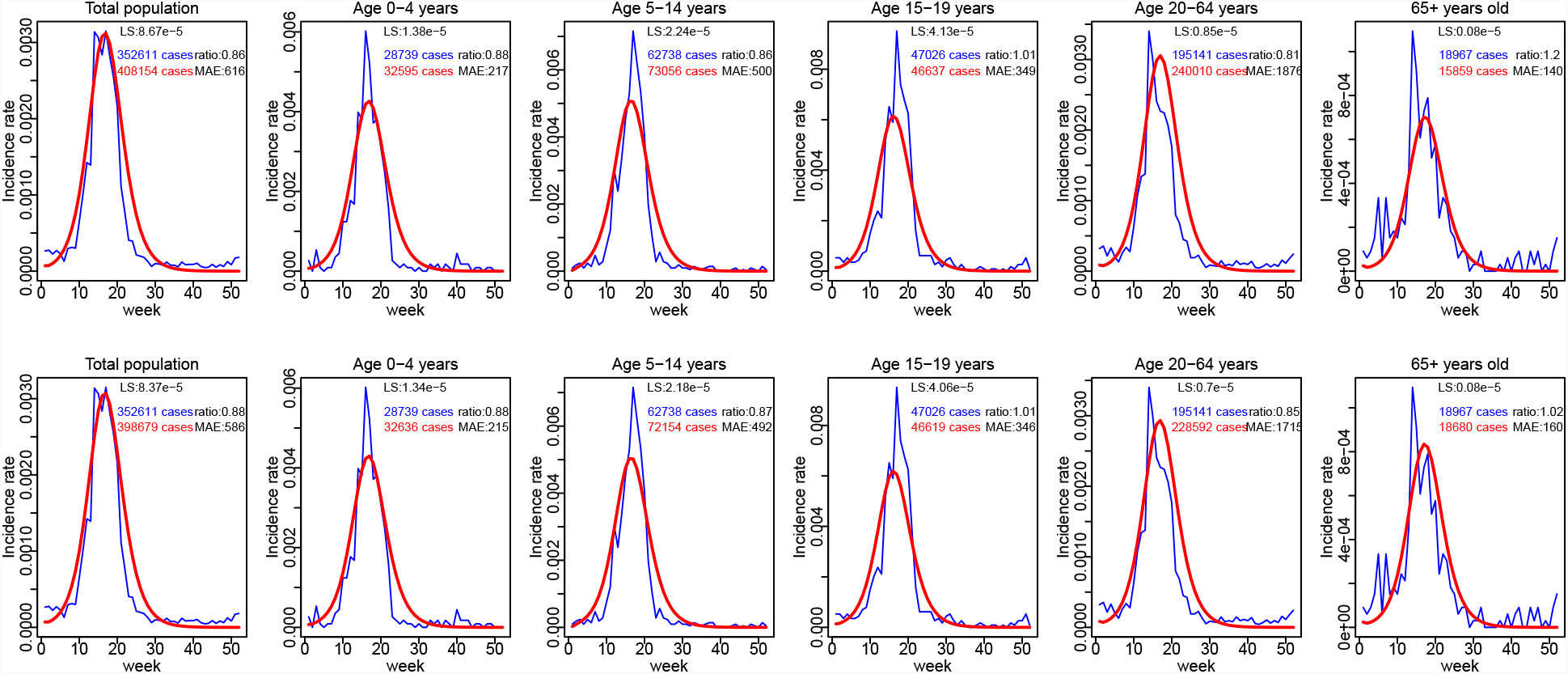
Observed ILI incidence rates in Belgium 2010-2011 (blue) and corresponding model-based estimates (red) using overall contact matrices (top row) or exposure time matrices (bottom row)

## Discussion and conclusions

We analysed time use and social contact data and compared their use as proxy for effective contacts governing disease transmission for disease transmitted through the respiratory route. Our data set is unique since it provides both time use and social contact data from the same participants, avoiding possible differences due to sample biases. In our analysis we identified the main drivers in shaping everyday time use and linked this info to social contact patterns.

The reported time use patterns in Belgium are quite similar to the published patterns for Italy [37], but different from the results from Zimbabwe [38]. In Zimbabwe, the working-age participants and children less than 6 years old reported much less time at work and school, respectively, compared with participants of the same age group in Belgium and Italy. We found that males spent on average ±2 hours more at work than females, which is in line with previous work [39, 40]. We also found that both males and females living with children spend more time at home than people living without children, which is consistent with what was found in [39]. The expected temporal patterns were observed, with more time spent at “other” locations during weekends and holiday periods. The time spent at home seemed not to be affected by the type of day. Our gender-specific analysis of the time use data indicated that participants were prone to spend more time with the same gender when they are young, and more time with the other gender when they are older. This result differs from the observed gender-specific contact rates as reported in [19], where the assortativity is reported to be higher for same-gender contacts.

Power law dispersal has been useful to model disease counts [13, 14, 15], though questions remained whether this also holds for social contact behavior, the driver of transmission dynamics [2]. Danon et al. [16] showed a relation between clustering and distance from home, with high clustering within two miles, dominated by home contacts, but the highest value of clustering occurring at a distance 50 miles or more from home. The authors hypothesize that this might be due to differences in the purpose behind contacts made at various distances.

In our study, we observed an increase in the number of contacts by distance during weekdays until an age-specific distance-threshold. A large-scale study in Taiwan [41] reported that 52.7% of contacts took place at a distance less than 1 km from home, 29.2% at a distance 1-9 km from home, 14.6% at a distance 10-49 km from home. In our study, this pattern was clearly age-specific. Half of our reported contacts during regular weekdays for children between [0-18) years of age took place less than 1 km from home. During weekends, we observed more contacts at +10 km from home. In general, we observed that the average number of contacts decreased by distance, while the individual-level pattern is reverse. As such, social contact patterns conditional upon presence at each distance provided useful info to inform spatial transmission models. A study in China quantified the distances from home based on the latitude and longitude of each reported contact and observed an increase in assortative mixing when contacts were made further from home [42]. We observed similar patterns with the construction of conditional social contact matrices by distance. A study in the United Kingdom [43] requested for infants to report the maximum distance travelled from home, but, to date, did not report results thereof. Other survey designs (e.g., [38]) included the distance between home and work but did not report results related to this information, yet.

The age-specific social contact and time use patterns followed a similar trend of assortativeness, which stresses once more the tendency for people to interact with people of a similar age. In addition to that, strong mixing among generations (parent-child) was present in both the social contact and time use matrices. The inter-generation mixing is mostly observed at home, and was more pronounced in the exposure time matrix than in the contact matrix. With our unified survey, we can confirm the contrasting effect of the relative small number of social contacts at home compared to the large amount of time spent at home, as observed in [10, 38].

Social contact matrices provide useful data to estimate disease transmission dynamics in terms of the transmission dynamics and relative incidence [1, 2]. As such, we estimated the R_0_ for each distance conditional upon presence, and observed a clear increase by distance. This reflects an increasing transmission potential by distance from the individual-level perspective. The latter is of interest for meta-population and individual-based models, were individuals join other sub-populations at distance. We found that a constant (or decreased) social mixing behavior by distance conditional upon presence is likely to underestimate the transmission potential. Some individual-based models handled this by the use of location-specific mixing patterns irrespective of distance from home [44].

We compared the value of different social contact features (duration, physical/non-physical, etc.) to inform transmission models for parvovirus-B19 and VZV. By scoring the model-based prevalence with Belgian serological data, we found that physical contacts provided the best proxy for both parvovirus-B19 and VZV. In terms of contact duration, the best model fit was obtained with physical contacts of long duration (more than 4 hours) for parvovirus-B19 and (more than 1 hour) for VZV. Goeyvaerts et al. [3] reported the best fit to VZV with physical contacts of at least 15 minutes; this result is also in line with the study of [4]. However, the previous studies did not analysed all combinations in terms of contact duration and physical/non-physical contacts, which explains the new “best estimate” in our study.

We also compared the results of the contact approach, the time use approach and the suitable contact approach in fitting serological data of VZV and parvovirus-B19. In the case of VZV, the suitable contact approach provided the best fit, while for parvovirus-B19, the time use approach gave the best fit. Our results are consistent with the findings of De Cao et al. [11], although we observed much higher parameter estimates for *q*_2_ (0.13 vs for parvovirus-B19 and 0.94 vs 0.37 for VZV), but the confidence intervals of these parameter estimates were overlapping.

We also tested the value of contact matrices and exposure time matrices in fitting the dynamic transmission model to weekly ILI incidence data in the season 2010-2011 in Belgium. Exposure time matrices provided a better fit to ILI than overall contact matrices and the integration of these two types of matrices did not improve the fit to the data. Within the social contact approach, physical contacts provided a better proxy for the risks of influenza transmission than non-physical contacts. We found that *R*_0_ of the best model is 1.43, this result is consistent with a systematic review of estimates of *R*_0_ for different types of influenza [45] and a study in UK [35] that also used contact matrices to fit to ILI incidence data. However, this value is lower than the estimates in [34], in which physical contact matrices from the Belgian POLYMOD data were used to fit to ILI data over multiple influenza season from 2003 to 2009. The difference in *R*_0_ can be explained by the relatively low number of ILI cases reported in the 2010-2011 season we used in this study. In addition, note that small differences in model parametrization entail substantial differences between the estimates of *R*_0_, which was also mentioned in [34].

In our study, we combined information from time use and social contact data to gather information on human mixing patterns. One of the main advantages with respect to previous work is that both sources of information came from the same survey. To keep participants’ burden as low as possible, time use information was collected with rather large time slots and participants were asked to fill in only one location for each time slot. However, the comparison with more refined time use surveys performed in Flanders [39, 40] confirms that we were able to well characterize time use patterns at an aggregated level. Therefore, we expect that this limitation did not substantially affect our results.

Our work emphasises the value of the unique and complementary information from time use and social contact data for analysing behavior and informing disease transmission models. Depending on the disease, one of the two data sources may be a better proxy for the transmission route of the pathogen. In particular, combining social contact and time use data can provide an improved measure of risk events with respect to VZV. Furthermore, our analysis based on data from the same survey is in line with studies that merged information from different surveys [10, 11]. This indicates that complementing social contact with independent time use data is a viable choice for the analyses presented here.

## Data Availability

The social contact datasets analyzed during the current study are available on Zenodo. 
The seroprevalence data are available on the SIMID website. The ILI incidence data is available on the Epistat website

https://doi.org/10.5281/zenodo.4059825

https://www.simid.be/modeling-infectious-disease-parameters-based-on-serological-and-social-contact-data/

https://epistat.wiv-isp.be/influenza/

## Declarations

### Ethics approval and consent to participate

The contact dataset used for the current study are available online within the social contact data sharing initiative which is part of the ERC consolidator grant “TransMID” which received ethical approval from the Hasselt University Medical Ethical Committee (CME2016/618).

The seroprevalence data are anonymous and openly available on the SIMID website (https://www.simid.be/modelinfectious-disease-parameters-based-on-serological-and-social-contact-data/).

The Belgian National Reference Centre of Influenza (BELNIC) is responsible for the laboratory surveillance of influenza in Belgium. The ILI incidence data is anonymous and available on the Epistat website [46].

### Consent for publication

Not applicable.

## Availability of data and materials

The social contact datasets analysed during the current study are available on Zenodo [47]. The seroprevalence data are available on https://www.simid.be/modeling-infectious-disease-parameters-based-on-serological-and-social-contact-data/ and the ILI incidence data is available on [46].

## Author’s contributions

NH and PB designed and coordinated the survey. TVH and LW conceived the study and laid out a paper structure. LW and KVK performed data cleaning. TVH, LW and KVK performed the data analyses. TVH and LW drafted the manuscript in consultation with all the other authors. NH, PC, KVK, JM and PB made substantial revisions to the manuscript. All authors contributed to the final version of the manuscript. All authors read and approved the final manuscript.

## Acknowledgements

We greatly thank the Scientific Institute of Public health for their permission to use the ILI data.

## Figures

**Additional Files Additional file 1 Missing data exploration, descriptive analysis results and the list of variables included in the imputation model**.

**Additional file 2 Time use at different locations by gender and age**.

**Additional file 3 The mean daily exposure time among age groups across locations**.

**Additional file 4 Number of contacts and temporal social contact matrices by distance**.

**Additional file 5 Fitting results of social contact and exposure matrices to parvovirus-B19 and VZV serological data and ILI incidence data**.

