## Supplementary files for "Exploring human mixing patterns based on time use and social contact data and their implications for infectious disease transmission models"

### Additional file 1

#### Missing data exploration, descriptive analysis results and the list of variables included in the imputation model

Table S1: Missing frequency in time use data

| Nr of not-filledin | 0 | 1 | 2 | 3 | 4 | 5 | 6 | 7 | 8 | 9 | 10 | 11 | 12 | 13 | 14 | 15 | 16 | 17 |
|---|---|---|---|---|---|---|---|---|---|---|---|---|---|---|---|---|---|---|
| Nr of participants | 1576 | 14 | 12 | 10 | 8 | 4 | 5 | 3 | 6 | 3 | 3 | 1 | 2 | 1 | 1 | 6 | 3 | 49 |

Table S2: Missing frequency by diary versions. Version 1 is for children less than 13, version 2 is for participants from 13 to 65 and version 3 for participants older than 65.

| Nr of not-filledin | Version 1 | Version 2 | Version 3 |
|---|---|---|---|
| 0 | 283 | 1012 | 281 |
| 1+ | 13 | 90 | 28 |

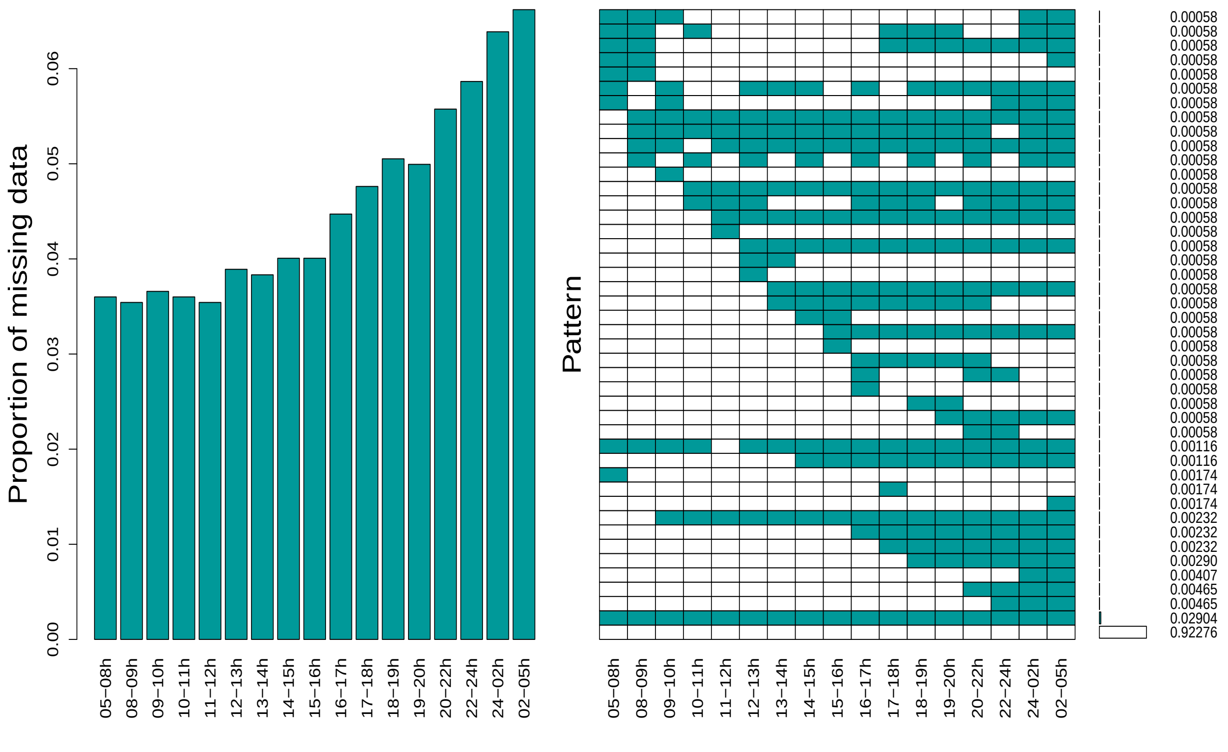


Figure S1: Missing data by time slots

#### Descriptive analysis

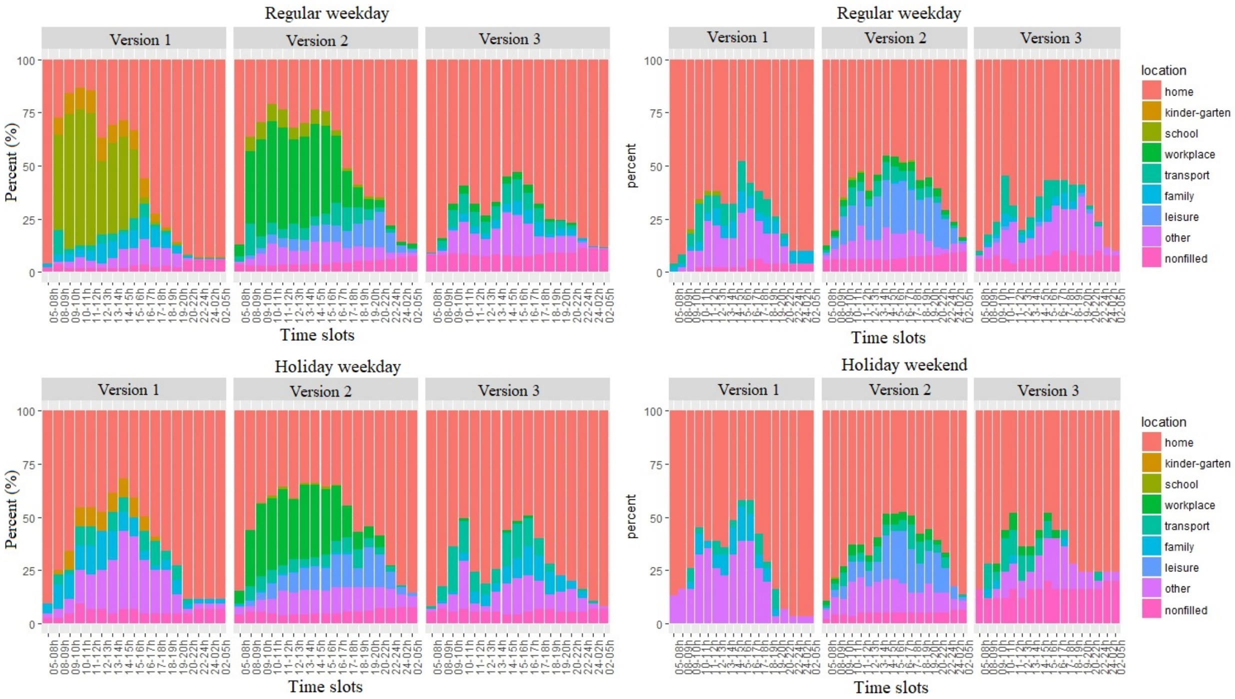


Figure S2: Time spent at different locations, temporal factors and diary versions

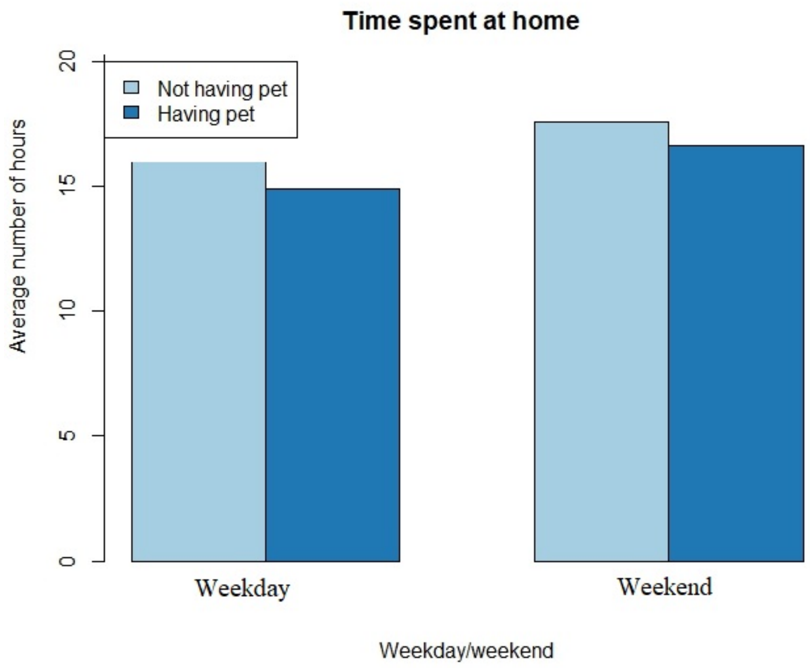


Figure S3: Owning animal and time use at home

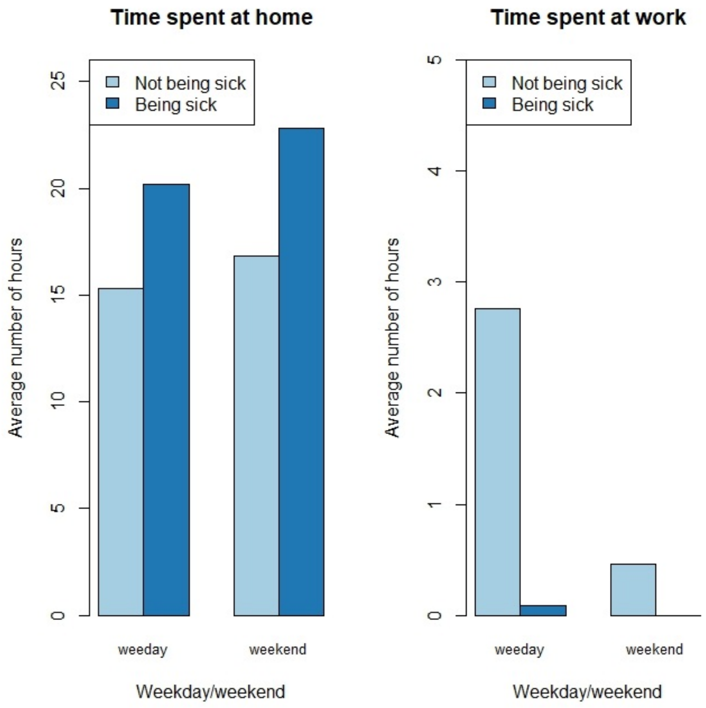


Figure S4: Time use when being ill and being healthy

#### Variables included in the imputation model

Table S3: Variables included in the imputation model

| Nr | Variable name | Variable labels | Type | Value labels |
|---|---|---|---|---|
| 1. | participant_age | Age of survey participants | Integer | |
| 2. | participant_gender | Gender of survey participants | String 1 char | M: male<br>F: female |
| 3. | dayofweek | Day of the week of filling in the diary | Integer | 0: Sunday<br>1: Monday<br>2: Tuesday<br>3: Wednesday<br>4: Thursday<br>5: Friday<br>6 : Saturday |
| 4. | holiday | Was the diary filled in during a public or school holiday? | String 1 char | Y: Yes<br>N: No |
| 5. | time_use_location_1 | Location where you spent the most time between: 5-8h | Integer | 1: home<br>2: kinder-garden<br>3: school<br>4: workplace<br>5: transport<br>6: family<br>7: leisure<br>8: other<br>9: missing |
| 6. | time_use_location_2 | Location where you spent the most time between: 9-10h | Integer | As above |
| 7. | time_use_location_3 | Location where you spent the most time between: 10-11h | Integer | As above |
| 8. | time_use_location_4 | Location where you spent the most time between: 11-12h | Integer | As above |
| 9. | time_use_location_5 | Location where you spent the most time between: 12-13h | Integer | As above |
| 10. | time_use_location_6 | Location where you spent the most time between: 13-14h | Integer | As above |
| 11. | time_use_location_7 | Location where you spent the most time between: 14-15h | Integer | As above |
| 12. | time_use_location_8 | Location where you spent the most time between: 15-16h | Integer | As above |
| 13. | time_use_location_9 | Location where you spent the most time between: 16-17h | Integer | As above |
| 14. | time_use_location_10 | Location where you spent the most time between: 17-18h | Integer | As above |
| 15. | time_use_location_11 | Location where you spent the most time between: 18-19h | Integer | As above |
| 16. | time_use_location_12 | Location where you spent the most time between: 19-20h | Integer | As above |
| 17. | time_use_location_13 | Location where you spent the most time between: 20-22h | Integer | As above |
| 18. | time_use_location_14 | Location where you spent the most time between: 22-24h | Integer | As above |
| 19. | time_use_location_15 | Location where you spent the most time between: 24-02h | Integer | As above |
| 20. | time_use_location_16 | Location where you spent the most time between: 02-05h | Integer | As above |
| 21. | time_use_location_17 | Location where you spent the most time between: 05-08h | Integer | As above |
| 22. | hh_size | Household size including participants | Integer | |

#### Calculation of Confidence Interval (CI)

MI is used for the dataset $\mathcal{D} = \{\mathcal{D}^{obs}, \mathcal{D}^{mis}\}$ where the data matrix consist of both observed and missing values. For each of the $\mathcal{M}$ imputed datasets $\mathcal{D}_m$, $\mathcal{B}$ bootstrap samples are drawn which yields $\mathcal{M}$ x $\mathcal{B}$ datasets $\mathcal{D}^*_{m,b}$ where $b = 1, ...., \mathcal{B}$ and $m = 1, ...., \mathcal{M}$. In each of these datasets, the parameter of interest is estimated, $\hat{\theta}^*_{m,b}$. The pooled set of ordered estimates $\Theta^*_{MB} = \{\hat{\theta}^*_{m,b}; b = 1, .., \mathcal{B}; m = 1, ...., \mathcal{M}\}$ is used to construct the 1-2$\alpha$ % confidence interval for $\theta$ [4]:

$$[\hat{\theta}_{lower}; \hat{\theta}_{upper}] = [\hat{\theta}^{*,\alpha}_{MB}; \hat{\theta}^{*,1-\alpha}_{MB}] \quad (1)$$

Where $\hat{\theta}^{*,\alpha}_{MB}$ is the $\alpha$ percentile of the ordered estimates $\Theta^*_{MB}$

### Additional file 2

#### Time use at different locations by gender and age

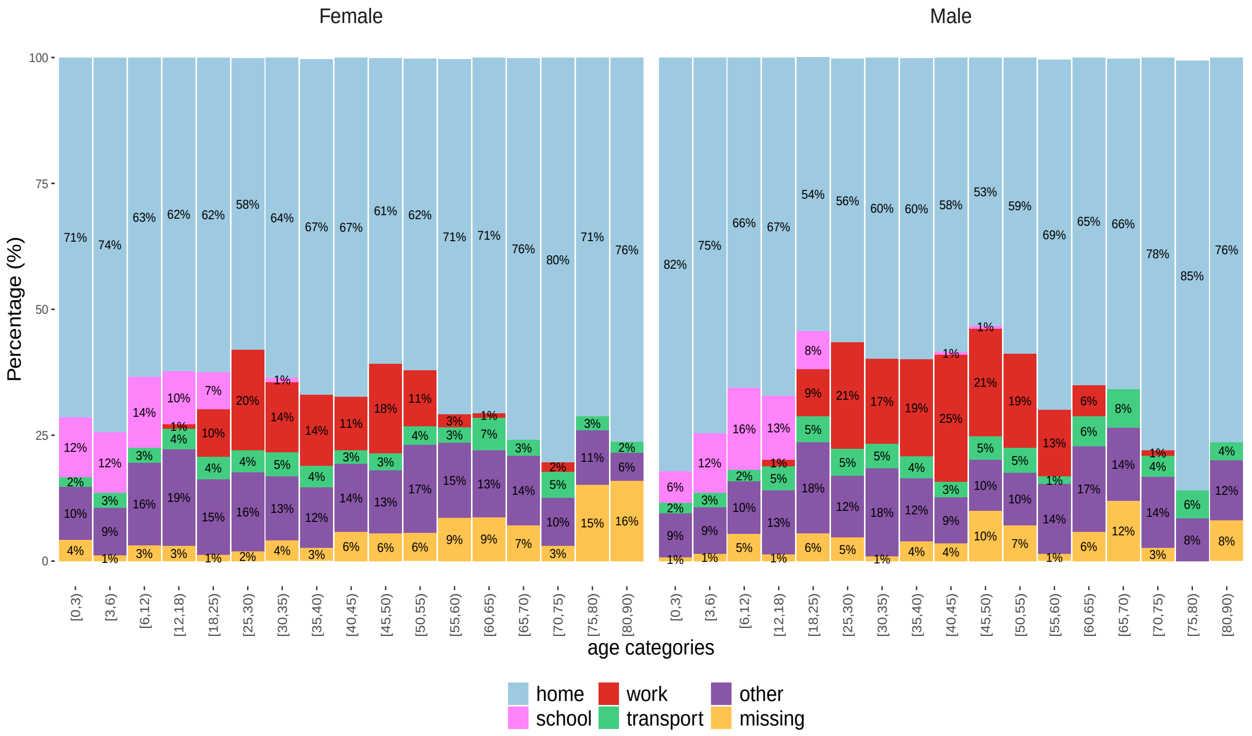


Figure S1: Time use per day over age categories of females and males (values below 1% are not shown)

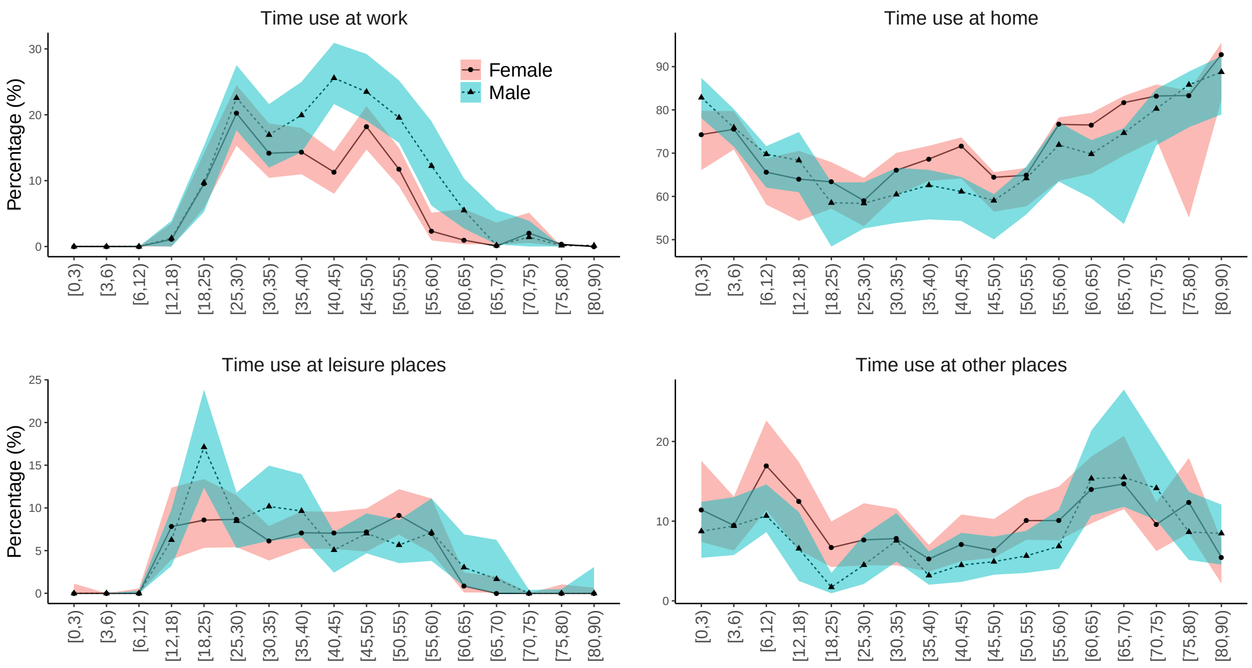


Figure S2: Time use of the male and the female over age groups

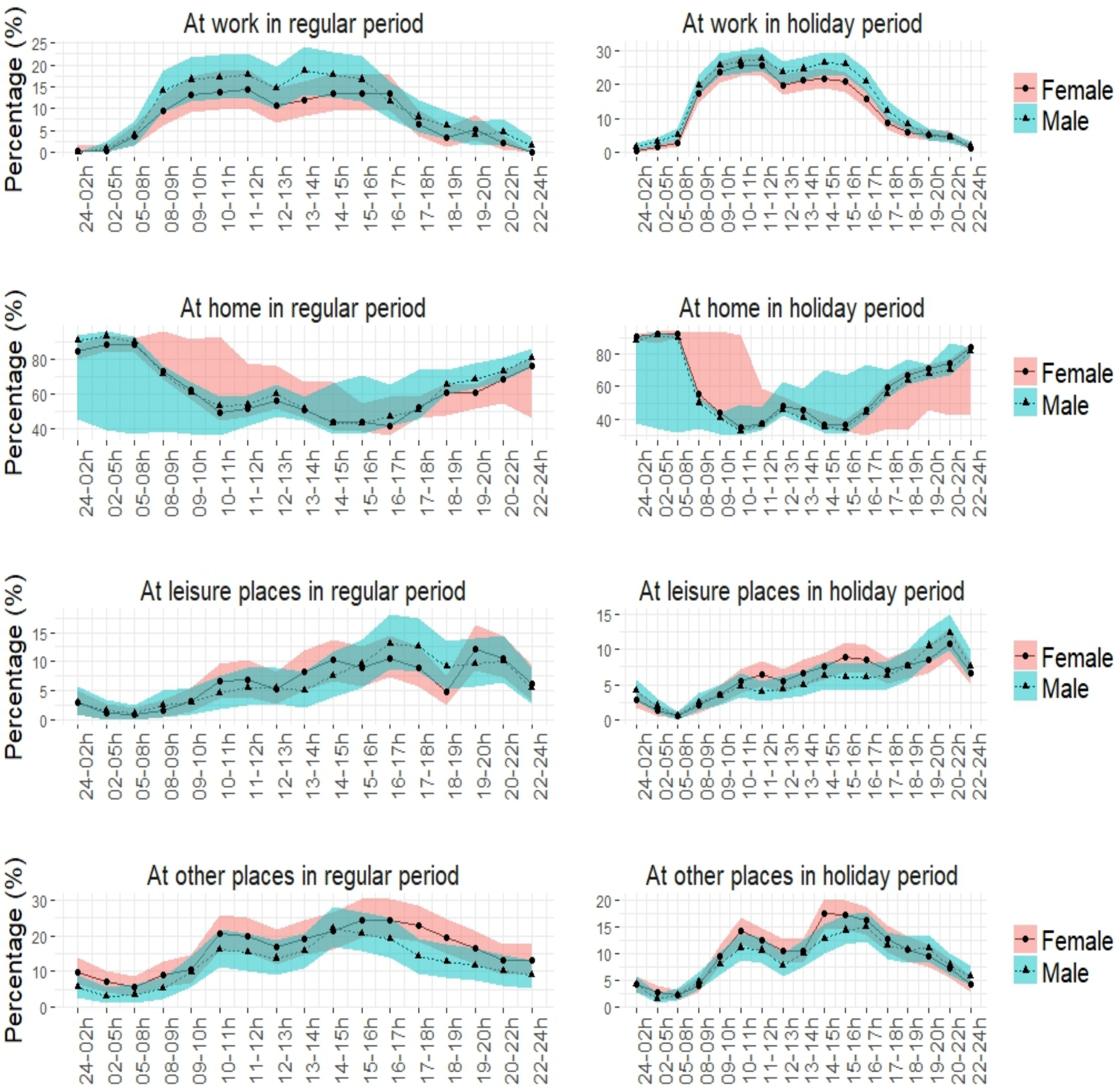


Figure S3: Time use of the female and the male at work, home, leisure and other places over time slots during a day in regular and holiday period

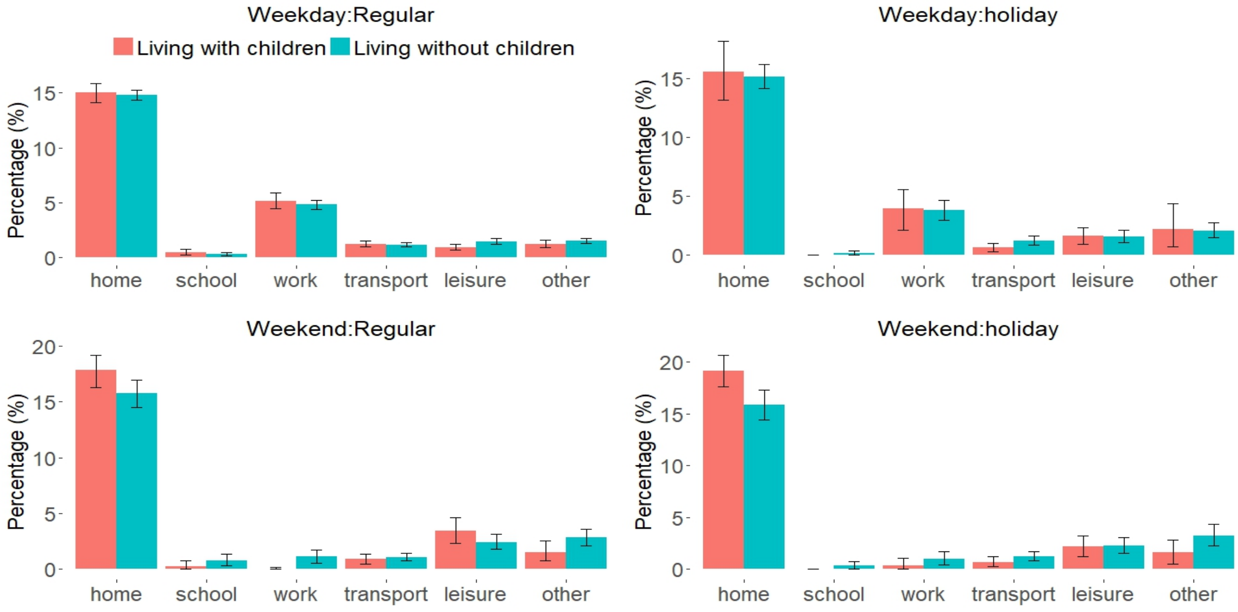


Figure S4: Time use per day by family situations (population from 25 to 65 years of age)

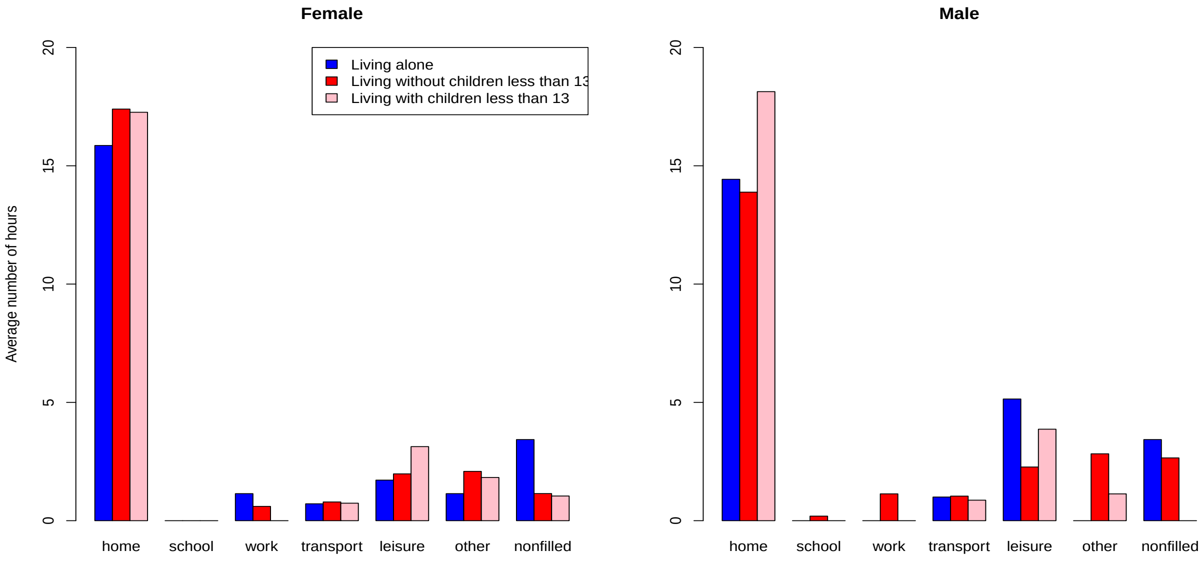


Figure S5: Time use per day by family characteristics in holiday period (population from 25 to 65 years of age)

### Additional file 3

#### Kullback divergence method

The compositional data, a special type of multivariate data, is illustrated as below:

$$S^d = \left\{ (y_1, ...., y_D | y_i \geq= 0), \sum_{i=1}^{D} y_i = 1 \right\} (d = D - 1) \tag{2}$$

Kullback-Leibler divergence can express the dissimilarity between two probability distributions. In this case, it can be used for minimization of the distance between the observed and the fitted compositions with respect to the coefficients ([5]).

$$\sum_{j=1}^{n} \sum_{i=1}^{D} KL(y_{ij}, \hat{y}_{ij}) = \sum_{j=1}^{n} \sum_{i=1}^{D} KL(y_{ij}, f_{ij}\ (\beta; x)) = \sum_{j=1}^{n} \sum_{i=1}^{D} y_{ij} log \frac{y_{ij}}{f_{ij}\ (\beta; x)} \tag{3}$$

Where:

- D: number of response variables (better known as components). Each response variable is a proportion, summing up to 1, hence (D-1) variables included in the model.
- n: a total of observations
- $\hat{y}_{ij}$ is the $j^{th}$ predicted observation for $i^{th}$ composition.

#### Mean daily time of exposure

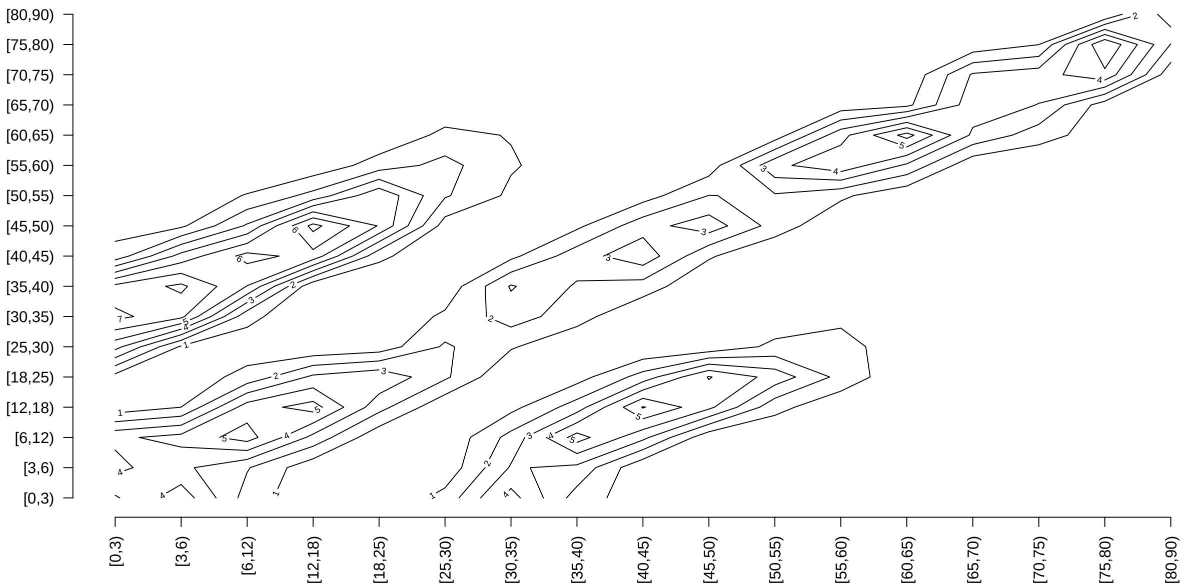


Figure S1: Mixing patterns by age based on reported presence and time spent at home

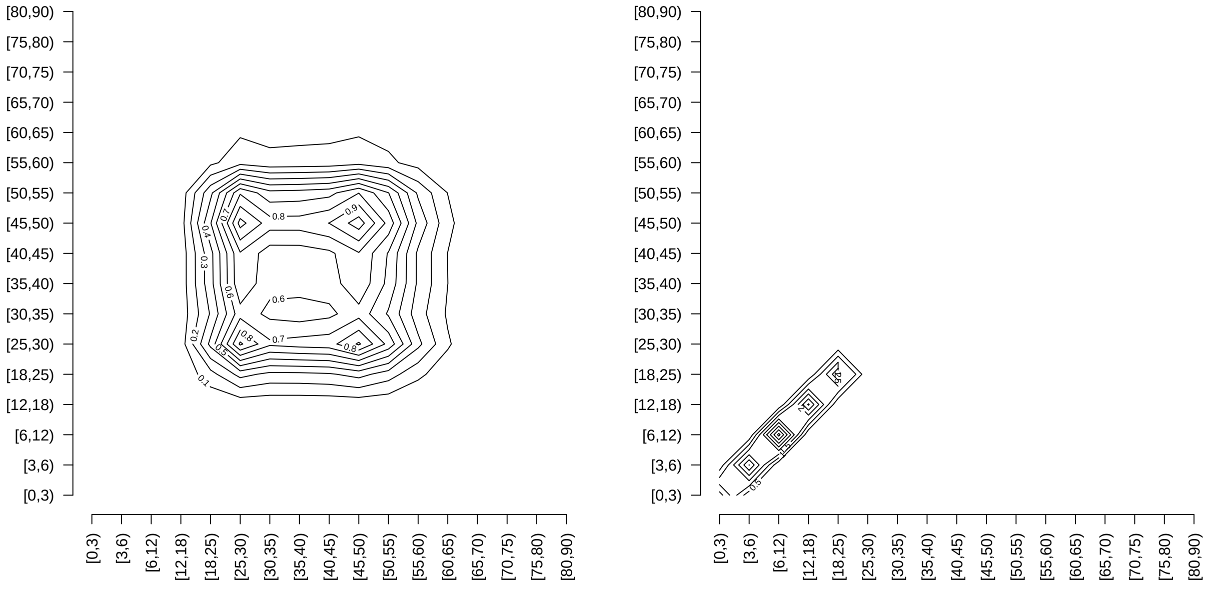


Figure S2: Mixing patterns by age based on reported presence and time spent at work (left) and school (right)

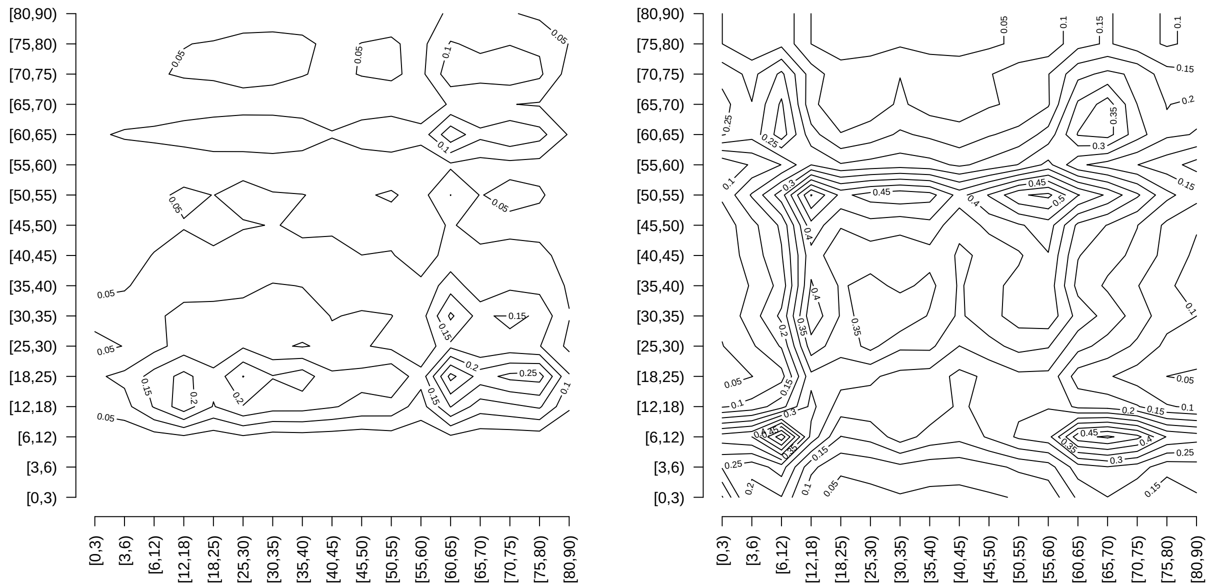


Figure S3: Mixing patterns by age based on reported presence and time spent at transport (left) and other locations (right)

Table S1: Mean daily time of exposure between people in age group i (in the rows) and people in age group j ( in the columns)

| | **[0;3)** | **[3;6)** | **[6;12)** | **[12;18)** | **[18;25)** | **[25;30)** | **[30;35)** | **[35;40)** | **[40;45)** | **[45;50)** | **[50;55)** | **[55;60)** | **[60;65)** | **[65;70)** | **[70;75)** | **[75;80)** | **[80;90)** |
|---|---|---|---|---|---|---|---|---|---|---|---|---|---|---|---|---|---|
| **[0;3)** | 4.16 | 2.19 | 2.04 | 0.17 | 0.13 | 1.58 | 5.12 | 2.39 | 0.25 | 0.14 | 0.13 | 0.08 | 0.35 | 0.30 | 0.22 | 0.15 | 0.19 |
| **[3;6)** | 2.28 | 5.00 | 2.02 | 0.33 | 0.21 | 0.25 | 2.99 | 2.86 | 0.81 | 0.24 | 0.24 | 0.17 | 0.33 | 0.27 | 0.28 | 0.18 | 0.20 |
| **[6;12)** | 1.14 | 1.08 | 11.05 | 2.05 | 0.36 | 0.24 | 0.88 | 3.00 | 3.25 | 0.88 | 0.39 | 0.24 | 0.38 | 0.32 | 0.30 | 0.21 | 0.20 |
| **[12;18)** | 0.09 | 0.17 | 1.98 | 9.12 | 1.96 | 0.52 | 0.70 | 0.92 | 4.39 | 4.28 | 1.23 | 0.34 | 0.33 | 0.19 | 0.21 | 0.13 | 0.10 |
| **[18;25)** | 0.06 | 0.08 | 0.27 | 1.53 | 6.28 | 1.49 | 0.87 | 0.90 | 1.58 | 4.56 | 3.77 | 0.82 | 0.39 | 0.15 | 0.18 | 0.10 | 0.10 |
| **[25;30)** | 0.87 | 0.13 | 0.24 | 0.53 | 1.93 | 3.56 | 1.91 | 1.37 | 1.33 | 1.84 | 2.46 | 1.30 | 0.65 | 0.18 | 0.22 | 0.11 | 0.09 |
| **[30;35)** | 2.70 | 1.51 | 0.83 | 0.68 | 1.09 | 1.84 | 3.01 | 2.51 | 1.28 | 1.37 | 1.42 | 0.81 | 0.61 | 0.21 | 0.24 | 0.13 | 0.10 |
| **[35;40)** | 1.27 | 1.45 | 2.85 | 0.91 | 1.14 | 1.32 | 2.52 | 2.75 | 2.25 | 1.55 | 1.23 | 0.57 | 0.51 | 0.15 | 0.22 | 0.13 | 0.11 |
| **[40;45)** | 0.12 | 0.38 | 2.86 | 4.00 | 1.85 | 1.19 | 1.19 | 2.09 | 4.33 | 2.42 | 1.24 | 0.50 | 0.38 | 0.21 | 0.23 | 0.13 | 0.07 |
| **[45;50)** | 0.07 | 0.11 | 0.74 | 3.72 | 5.07 | 1.58 | 1.22 | 1.37 | 2.31 | 5.28 | 2.27 | 0.60 | 0.35 | 0.13 | 0.24 | 0.11 | 0.08 |
| **[50;55)** | 0.06 | 0.12 | 0.35 | 1.14 | 4.50 | 2.25 | 1.36 | 1.16 | 1.27 | 2.43 | 3.48 | 1.55 | 0.58 | 0.21 | 0.23 | 0.15 | 0.28 |
| **[55;60)** | 0.05 | 0.09 | 0.24 | 0.36 | 1.10 | 1.34 | 0.87 | 0.61 | 0.58 | 0.73 | 1.75 | 3.43 | 1.34 | 0.18 | 0.18 | 0.11 | 0.07 |
| **[60;65)** | 0.21 | 0.20 | 0.42 | 0.38 | 0.57 | 0.74 | 0.72 | 0.59 | 0.47 | 0.47 | 0.71 | 1.47 | 5.88 | 1.03 | 0.51 | 0.26 | 0.22 |
| **[65;70)** | 0.24 | 0.20 | 0.45 | 0.28 | 0.27 | 0.26 | 0.32 | 0.22 | 0.33 | 0.23 | 0.33 | 0.25 | 1.31 | 3.16 | 1.31 | 0.29 | 0.23 |
| **[70;75)** | 0.18 | 0.22 | 0.45 | 0.32 | 0.37 | 0.35 | 0.39 | 0.35 | 0.41 | 0.43 | 0.39 | 0.27 | 0.70 | 1.40 | 2.76 | 1.38 | 0.25 |
| **[75;80)** | 0.14 | 0.17 | 0.37 | 0.23 | 0.22 | 0.19 | 0.24 | 0.24 | 0.25 | 0.22 | 0.29 | 0.18 | 0.40 | 0.35 | 1.54 | 2.71 | 0.59 |
| **[80;90)** | 0.15 | 0.15 | 0.28 | 0.15 | 0.20 | 0.13 | 0.16 | 0.17 | 0.11 | 0.14 | 0.45 | 0.10 | 0.29 | 0.23 | 0.24 | 0.50 | 3.53 |

#### Model comparison

Table S2: Model comparison

| **Model** | **AIC** |
|---|---|
| Zero adjusted Gama model | 64776.79 |
| Zero adjusted IG model | 58408.50 |
| Zero adjusted LN model | **56596.11** |
| Zero adjusted WEI model | 62034.31 |
| Zero adjusted PARETO model | Not converged |

#### The proportion of the number of contacts and time use at different location over time settings

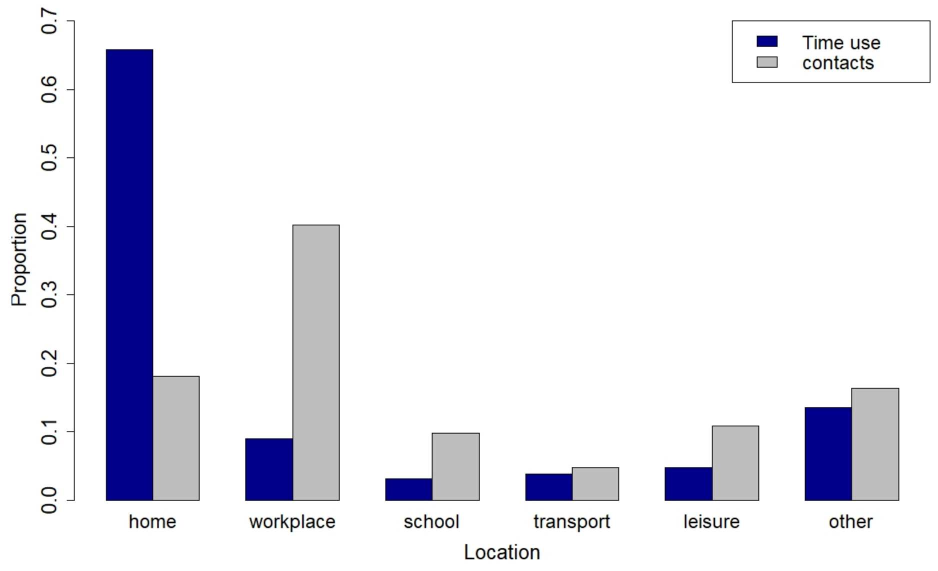


Figure S4: Proportion of number of contacts and time use

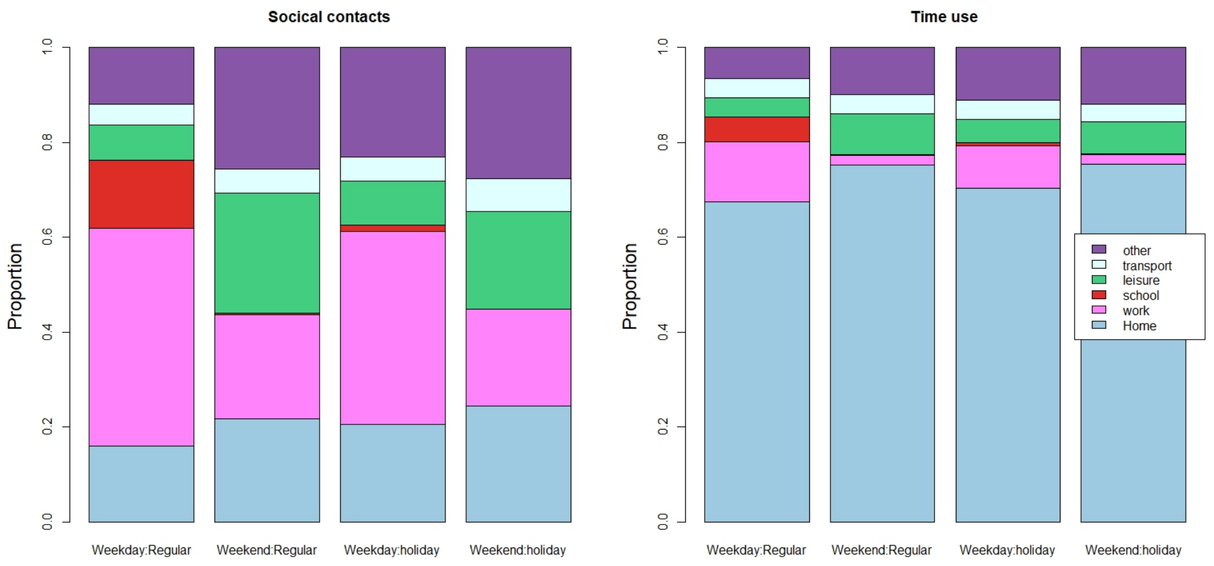


Figure S5: Proportion of number of contacts and time spent at each location in different types of day

### Additional file 4

#### Number of contacts and temporal social contact matrices by distance

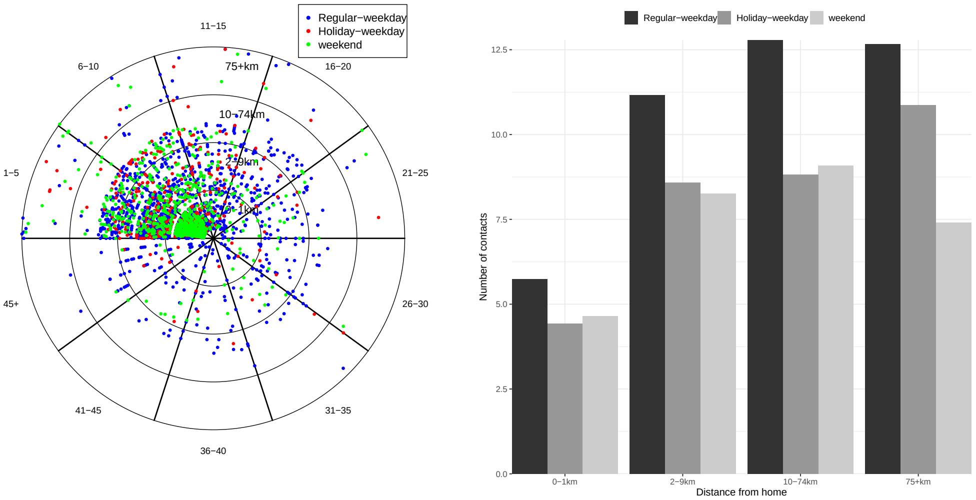


Figure S1: The number of overall contacts over distance upon presence

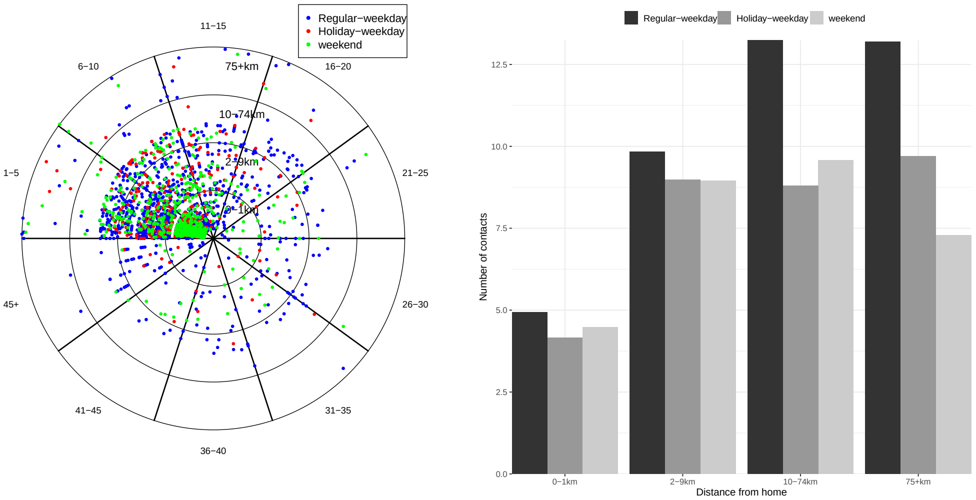


Figure S2: The number of overall contacts of adults over distance upon presence

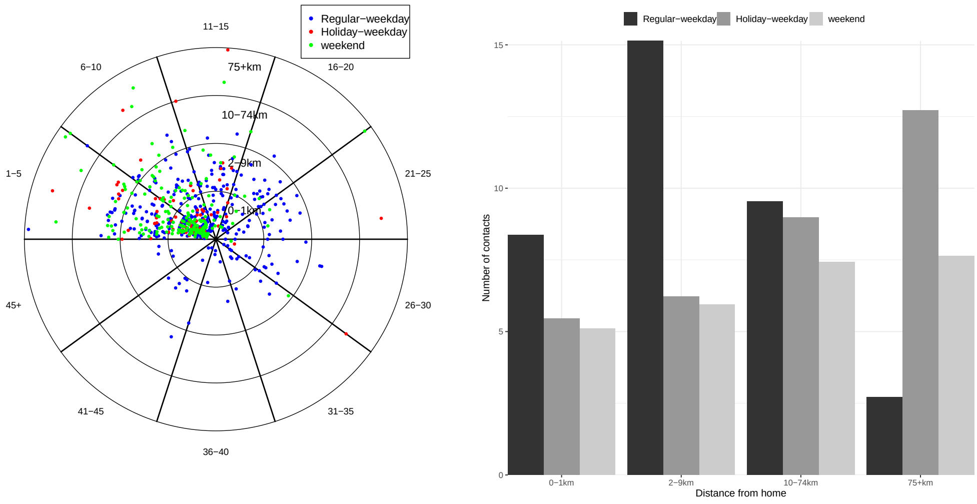


Figure S3: The number of overall contacts of children over distance upon presence

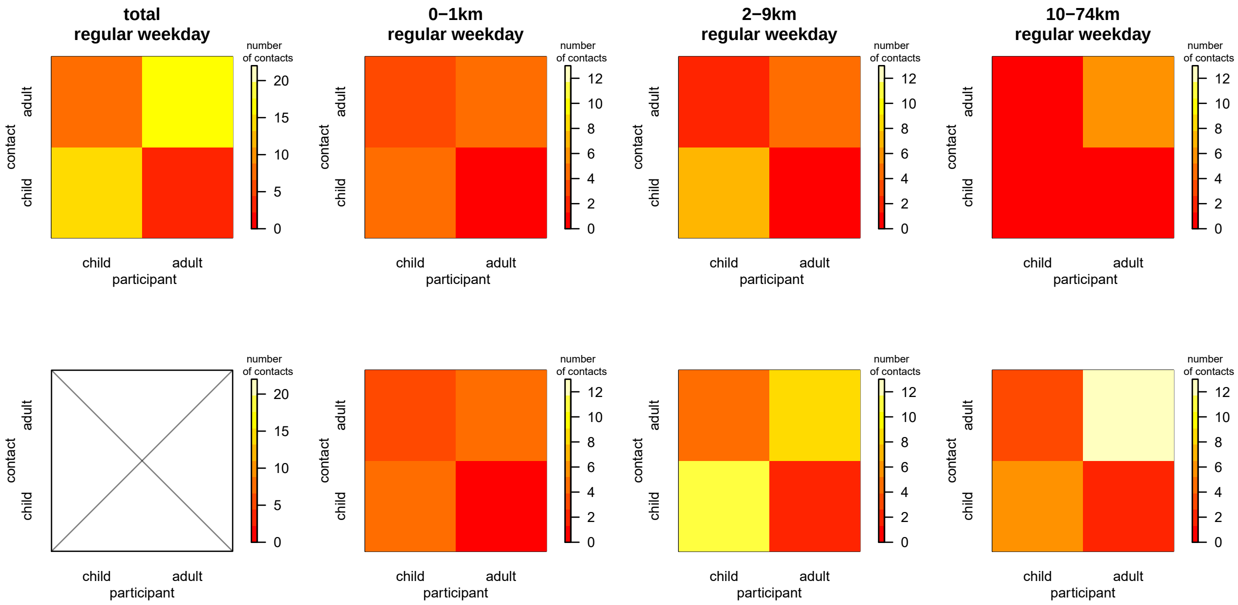


Figure S4: Social contact matrices on regular weekdays by distance, unconditional (top) and conditional (bottom) upon presence.

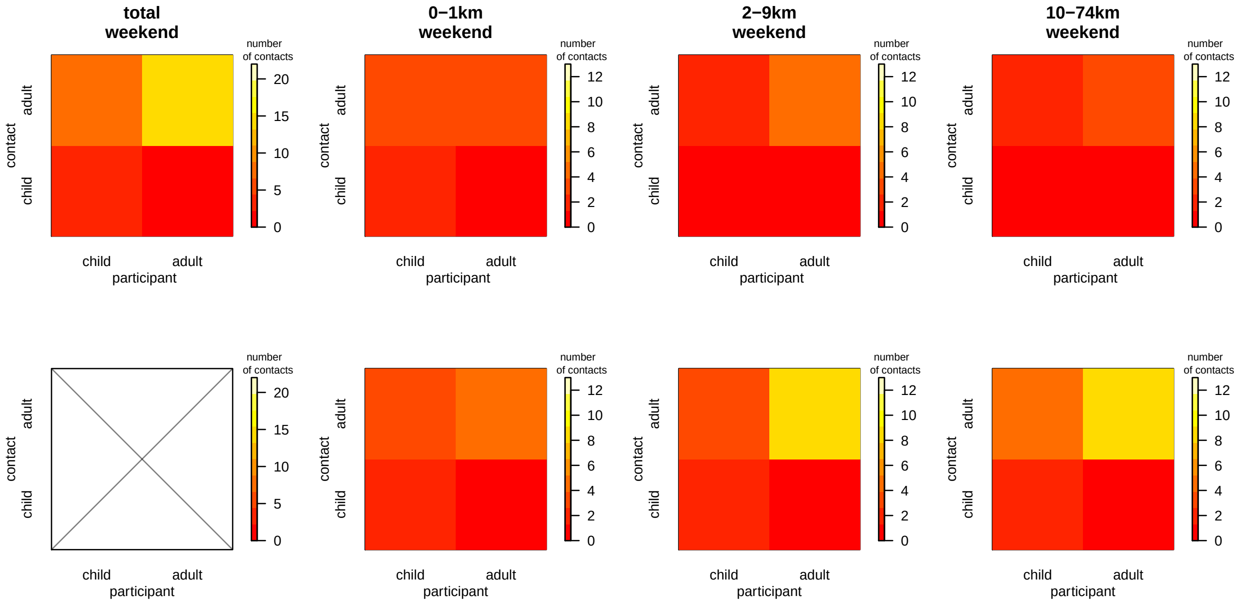


Figure S5: Social contact matrices on weekend days by distance, unconditional (top) and conditional (bottom) upon presence.

### Additional file 5

#### Model fit to serologial data

Table S1: MSIRWb model fits to parvovirus-B19 based on different matrices

| **Contact data** | $q_1$ | **95% CI for $q_1$** | $\epsilon$ | **95% CI of $\epsilon$** | $R_0$ | **95% CI of $R_0$** | **AIC** | $\Delta_{AIC}$ | **Weight** | **LER** |
|---|---|---|---|---|---|---|---|---|---|---|
| All contacts | 0.012 | [0.011;0.014] | 0.006 | [0.004;0.007] | 2.392 | [2.088;2.685] | 3473.016 | 3.856 | 0.039 | 0.838 |
| Close contacts | 0.022 | [0.019;0.024] | 0.006 | [0.004;0.007] | 2.025 | [1.827;2.207] | 3470.934 | 1.774 | 0.111 | 0.386 |
| Non-close contacts | 0.031 | [0.025;0.038] | 0.006 | [0.003;0.010] | 3.167 | [2.579;3.871] | 3481.550 | 12.390 | 5.5E-04 | 2.691 |
| Close < 15p | 0.153 | [0.096;0.232] | 0.005 | [0.005E-1;0.012] | 2.130 | [1.519;3.283] | 3496.605 | 27.445 | 3.0E-07 | 5.960 |
| Close > 15p | 0.025 | [0.022;0.029] | 0.006 | [0.004;0.007] | 2.011 | [1.835;2.197] | 3469.990 | 0.830 | 0.179 | 0.181 |
| Close > 1h | 0.029 | [0.025;0.033] | 0.005 | [0.004;0.006] | 1.880 | [1.743;2.006] | 3469.976 | 0.816 | 0.180 | 0.178 |
| Close > 4h | 0.056 | [0.0453;0.064] | 0.006 | [0.005;0.007] | 1.895 | [1.717;2.103] | **3469.158** | **0** | 0.271 | **0** |
| Linear combination of 5 locations | * | * | 0.019 | * | 1.675 | [1.552;3.401] | 3476.990 | 7.830 | 0.005 | 1.701 |
| **Time use data** | | | | | | | | | | |
| Overall exposure time | 0.010 | [0.008;0.011] | 0.007 | [0.006;0.008] | 2.075 | [1.905;2.301] | 3471.015 | 1.858 | 0.107 | 0.403 |
| Linear combination of 5 locations | * | * | * | * | 2.084 | [1.835;2.917] | 3472.352 | 3.194 | 0.055 | 0.694 |
| **Combined data** | | | | | | | | | | |
| | 0.097 | [0.019;0.250] | | | | | | | | |
| Suitable contacts | 0.129 | [0.037;0.925] | 0.007 | [0.006;0.008] | 2.152 | [1.995;2.296] | 3472.45 | 3.286 | 0.052 | 0.714 |

$q_1$ proportionality factor; [$q_2$] the fraction of total exposure time suitable for transmission in combined data; $\epsilon$: waning rate
* See Appendix 6 Table 1; $\Delta_{AIC}$: AIC difference; ER: Evidence ratio; LER: $log_{10}(ER)$.

Table S2: MSIR model fits to VZV based on different matrices

| **Contact data** | $q_1$ | **95% CI of $q_1$** | $R_0$ | **95% CI of $R_0$** | **AIC** | $\Delta_{AIC}$ | **Weight** | **LER** |
|---|---|---|---|---|---|---|---|---|
| All contacts | 0.045 | [0.035;0.055] | 7.309 | [5.843;9.081] | 1419.546 | 38.413 | 2.19E-09 | 8.341 |
| Close contacts | 0.081 | [0.065;0.094] | 6.389 | [5.162;7.456] | 1390.056 | 8.923 | 0.006 | 1.938 |
| Non-close contacts | 0.102 | [0.081;0.188] | 8.895 | [7.134;16.526] | 1487.455 | 106.322 | 3.93E-24 | 23.088 |
| Close < 15p | 0.545 | [0.347;0.859] | 6.510 | [4.613;10.754] | 1399.230 | 18.097 | 5.66E-05 | 3.930 |
| Close > 15p | 0.097 | [0.078;0.113] | 6.531 | [5.290;7.692] | 1388.845 | 7.712 | 0.010 | 1.675 |
| Close > 1h | 0.113 | [0.091;0.132] | 6.228 | [5.114;7.392] | 1385.650 | 4.517 | 0.050 | 0.981 |
| Close > 4h | 0.219 | [0.168;0.265] | 6.341 | [4.907;8.003] | 1386.845 | 5.712 | 0.028 | 1.240 |
| Linear combination of 5 locations | * | * | 7.401 | [5.767;11.326] | 1383.369 | 2.236 | 0.157 | 0.486 |
| **Time use data** | | | | | | | | |
| Overall exposure time | 0.037 | [0.030;0.041] | 6.893 | [5.717;7.778] | 1384.452 | 3.319 | 0.092 | 0.721 |
| Linear combination of 5 locations | * | * | 19.714 | | 1383.140 | 2.007 | 0.176 | 0.436 |
| **Combined data** | | | | | | | | |
| Suitable contacts | 0.068 | [0.061; 0.647] | **7.813** | **[6.832;8.477]** | **1381.133** | **0** | **0.481** | **0** |
| | 0.938 | [0.083; 0.952] | | | | | | |

$q_1$ proportionality factor; $q_2$ the fraction of total exposure time suitable for transmission in combined data;
* See Appendix 6 Table 1; $\Delta_{AIC}$: AIC difference; ER: Evidence ratio; LER: $log_{10}(ER)$.

Table S3: Location-specific proportional factors in the MSIRWb and MSIR model

| | The MSIRWb model | The MSIR model |
|---|---|---|
| **Contact data** | **fits to B19** | **fits to VZV** |
| Home | 1.39E-11 | 1.10E-10 |
| school | 1.88E-02 | 4.07E-02 |
| Work | 2.22E-13 | 2.68E-10 |
| Transport | 1.78E-04 | 3.29E-01 |
| General public | 3.23E-13 | 1.66E-13 |
| **Time use data** | **fits to B19** | **fits to VZV** |
| Home | 1.48E-03 | 9.78E-13 |
| school | 1.09E-02 | 1.88E-01 |
| Work | 1.12E-06 | 6.89E-01 |
| Transport | 1.80E-05 | 1.19E-10 |
| General public | 1.77E-01 | 8.49E-02 |

#### Model fit to ILI incidence data

The model is described by the following set of differential equations:

$$\frac{dS_i}{dt} = -q_i S_i \sum_j C_{i,j} I_j$$

$$\frac{dE_i}{dt} = q_i S_i \sum_j C_{i,j} I_j - rE_i$$

$$\frac{dI_i}{dt} = rE_i - fI_i$$

$$\frac{dR_i}{dt} = fI_i$$

where:

- $S_i$, $E_i$, $I_i$ and $R_i$ are the number of susceptible, exposed, infected and recovered individuals respectively in age group i.
- $C_{i,j}$ is the contact rate matrices and $q_i$ is age- dependent proportional factor; the product $q_i C_{i,j}$ translates into the average daily per capita rate at which an individual of age i makes effective contact with a person of age j.
- r is the rate at which an individual in the exposed class enters the infectious class
- f is daily rate at which infectious individuals recover and become immune.

We referred to [3, 7] and took an average latent period of 1 day (r=1) and an average infectious period of 3.8 days (f=1/3.8). We assumed that vaccination took place at the beginning of the October, with the coverage being 0.066% for 0-14 years, 5.5% for 15-19 years, 20% for 20-64 years, and 60% for 65 years and older [1]. The vaccinated people were assumed to be become immune and moved out the susceptible class immediately. The newly infected cases of the first week of the season 2010-2011, reported by GPs network [2, 6], were seeded in the model at time t=0. Given GPs network covering 1.75% of the population, we estimated infected cases from observed ILI data for the whole Belgian population. We estimated the model parameters by minimizing the sum of squared difference between the observed ILI incidence rate and the scaled model-based incidence rate, as presented in the following formula with i being age groups (0-4, 5-14, 15-19, 20-64 and $\geq 65$) and t is the index of week from 1 to 52 in the season 2010-2011. $\alpha$ is a scaling factor to account for several issues, e.g. consultation rate at GPs, mis-specification of model parameters...

$$\sum_i \sum_t \left[ \frac{I_{i,t}}{N_i} - \alpha \frac{\hat{I_{i,t}}}{N_i} \right]^2$$

The accuracy of the model is measured by the mean absolute error (MAE), where $I_t$ and $\hat{I}_t$ are observed number of cases and estimated number of cases at time t, respectively.

$$MAE = mean(\left| I_t - \hat{I}_t \right|)$$

Table S4: Model fits to ILI incidence data based on different matrices

| **Contact data** | $q_{1.1}$**(95% CI)** | $q_{1.2}$**(95% CI)** | $q_{1.3}$**(95% CI)** | $q_{1.4}$**(95% CI)** | $q_{1.5}$**(95% CI)** | $q_2$**(95% CI)** | **alpha(95%)** | **R0(95%)** | **LS value** |
|---|---|---|---|---|---|---|---|---|---|
| All contacts | 0.137(0.124-0.158) | 0.09 (0.079-0.098) | 0.131(0.114-0.148) | 0.090(0.084-0.096) | 0.120(0.086-0.122) | | 0.265(0.265-0.268) | 2.077(1.881-2.470) | 8.66e-5 |
| Non-close contacts | 0.420(0.326-0.546) | 0.207(0.198-0.394) | 0.257(0.149-0.451) | 0.165(0.0910-.329) | 0.005(0.003-0.009) | | 0.269(0.261-0.270) | 3.471(2.130-3.674) | 9.32e-5 |
| Close contacts | 0.212(0.162-0.325) | 0.167(0.104-0.213) | 0.274(0.210-0.421) | 0.204(0.125-0.381) | 0.242(0.210-0.391) | | 0.265(0.201-0.269) | 1.510(1.458-1.637) | 8.47e-5 |
| Close > 15p | 0.254(0.135-0.342) | 0.192(0.124-0.236) | 0.327(0.213-0.548) | 0.250(0.215-0j.451) | 0.301(0.281-0.428) | | 0.265(0.260-0.268) | 1.512(1.457-1.742) | 8.38e-5 |
| Close > 1h | 0.303(0.254-0.465) | 0.230(0.201-0.342) | 0.422(0.327-0.479) | 0.332(0.312-0.541) | 0.293(0.213-0.415) | | 0.266(0.262-0.269) | 1.429(1.324-1.543) | 8.28e-5 |
| Close > 4h | 0.544(0.326-0.642) | 0.438(0.218-0.526) | 0.807(0.231-0.902) | 0.673(0.429-0.791) | 0.514(0.491-0.615) | | 0.266(0.260-0.291) | 1.318(1.131-1.635) | 8.34e-5 |
| **Time use data** | | | | | | | | | |
| Overall exposure time | 0.097(0.081-0.102) | 0.076(0.069-0.812) | 0.119(0.107-0.129) | 0.08(0.077-0.082) | 0.149(0.94-0.198) | | 0.264(0.261-0.2661) | 1.630(1.401-1.722) | 8.37e-5 |
| **Combined data** | | | | | | | | | |
| Suitable contacts | 0.284(0.258-0.497) | 0.21(0.199-0.397) | 0.322(0.303-0.619) | 0.218(0.212-0.418) | 0.336(0.250-0.638) | 0.478(0.216-0.502) | 0.264(0.262-0.266) | 1.441(1.430-1.446) | 8.43e-5 |

$q_{1.1}$-$q_{1.5}$: The age-dependent proportionality factors;
$q_2$: The fraction of total exposure time suitable for transmission in combined data;
$\alpha$: the scaling factor

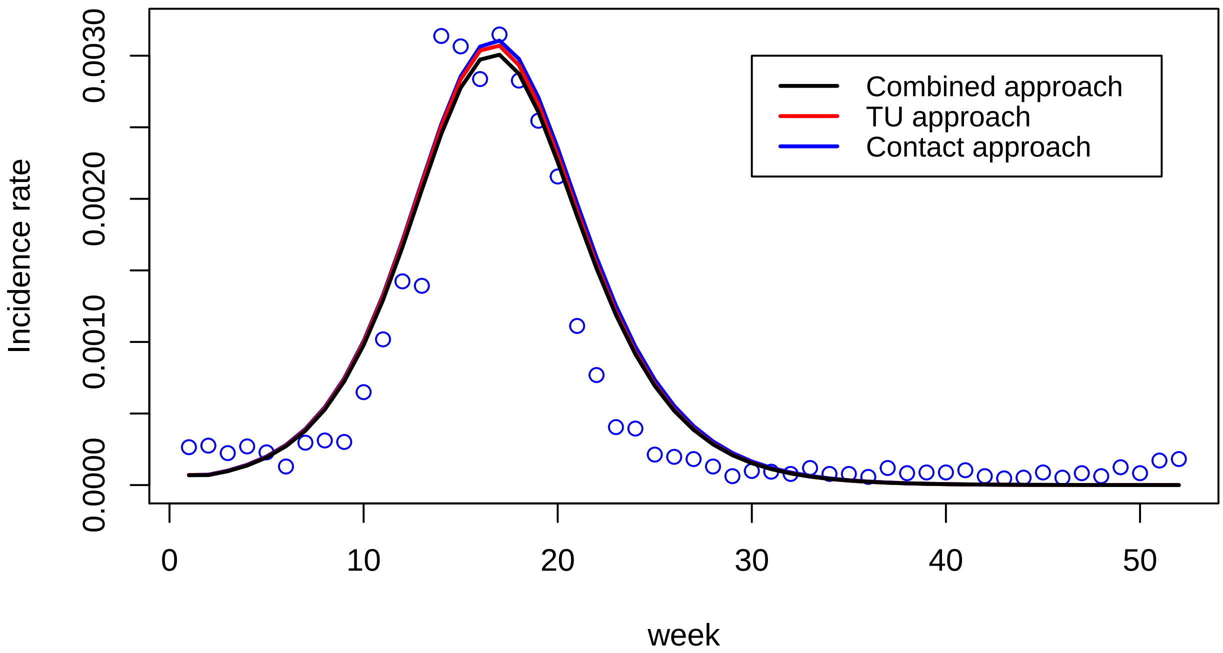


Figure S1: The fit to ILI incidence data.
